# Was CED the Right Choice? A Decision-Theoretic Evaluation of CMS’s ‘Cover with Evidence Development’ Policy for Aducanumab

**DOI:** 10.1101/2024.02.13.24302771

**Authors:** Jonah Popp, Eric Jutkowitz, Thomas Trikalinos

## Abstract

**Background:** In 2022, the Centers for Medicare & Medicaid Services (CMS) issued its final national coverage policy for aducanumab, a novel FDA-approved treatment for Alzheimer’s disease, deciding to ‘Cover with Evidence Development’ (CED). CMS will thus only pay for the treatment of AD patients enrolled in an approved randomized controlled trial (RCT). We sought to understand whether, given current evidence, CED was best from a societal perspective.

**Methods:** We conducted a modeling-based expected value of sample information analysis to estimate the expected net decision-theoretic value of a further RCT to evaluate the clinical efficacy of high-dose (10 mg/kg) aducanumab and to determine what sized trial, if any, is optimal conditional on an initial decision to cover or not. We also evaluated the expected net benefit of the manufacturer’s proposed RCT (‘ENVISION’). We considered two post-trial decision criteria: cost-effectiveness given updated evidence (‘efficiency’) and does the new trial demonstrate a statistical significant (p<0.05) clinical benefit. Results were used to calculate the expected population net monetary benefit (NMB) of four decision alternatives (including CED) depending on an initial coverage and trial decision. We ranked alternatives and calculated the expected opportunity loss of a suboptimal decision. We used a societal perspective and focused on willingness-to-pay (WTP) values for a quality-adjusted life year (QALY) between $50K-$200K. We conducted scenario analyses using different assumptions about population size, efficacy, and drug cost.

**Findings:** CMS’s decision to not cover aducanumab avoids an expected societal loss (NMB) of $15B-$110B. Even an optimally designed RCT would confer no or negative decision-theoretic value for WTP≤$100K or with statistical significance as a post-trial decision criterion, respectively, and thus denying coverage without a trial (rather than CED) is clearly preferable. For WTP=$150K (WTP=$200K) and assuming an efficiency criterion, CED with ENVISION or a similar trial is reasonable (decidedly optimal). The case for future research would become less ambiguous if the manufacturer again voluntarily dropped the price ≥50%.

**Interpretation:** The societal net value of a future trial (and thus CED) depends on how CMS would use the trial results to update its coverage decision and the WTP per QALY. Assuming CMS policymakers can avoid the pitfalls of a legal framework that limits their ability to consider costs in coverage decisions, the CED decision is at least reasonable, if not optimal, if a QALY is valued ≥$150K.

## Introduction

In June 2021, the US Food and Drug Administration (FDA) approved the human monoclonal antibody aducanumab to treat mild cognitive impairment (MCI) and mild Alzheimer’s Disease (AD).^1,2^ Aducanumab is the first FDA-approved treatment that purports to alter the clinical course of AD by clearing the accumulation of beta amyloid (Aβ) plaques.^2,3^ However, the FDA’s decision was controversial because aducanumab can cause edema and hemorrhage and its patient- and caregiver-relevant benefits are uncertain. The two existing identically-designed placebo-controlled randomized trials (RCTs), EMERGE and ENGAGE, were terminated early following a prespecified pooled futility analysis^2,4^ but had divergent results after additional data became available. Compared to placebo, a post-hoc analysis of available data from EMERGE showed a statistically-significant but modest and imprecisely estimated clinical benefit for high-dose recipients, while ENGAGE found no evidence of a benefit. Using trial results as input, a model-based cost-utility analysis by the Institute for Clinical and Economic Review (ICER) demonstrated that, given current evidence, the expected benefit of aducanumab does not justify the high costs^5^ under any plausible cost-effectiveness threshold.^6,7^

Because most MCI and mild AD patients are Medicare-eligible, the Centers for Medicare & Medicaid Services (CMS) would be the primary payer for aducanumab. In April of 2022, CMS issued its final national coverage policy for aducanumab^8^, deciding to ‘Cover with Evidence Development’ (CED) – by limiting reimbursement to beneficiaries enrolled in an FDA-approved or National Institute of Health (NIH) RCT.^8,9^ Aducanumab will consequently remain largely unavailable to patients in the near future. However, the CED policy incentivizes further research into aducanumab’s clinical efficacy and harm profile by committing to pay treatment costs for trial participants. Therefore, in the abstract, CED can be conceptualized as one of four general decision alternatives available to a centralized decision maker (Figure 1): (1) deny coverage and don’t conduct a (further efficacy) trial, (2) deny coverage and conduct a trial (CED), (3) cover without a trial, and (4) cover and conduct a trial.^10^

**Figure 1:**
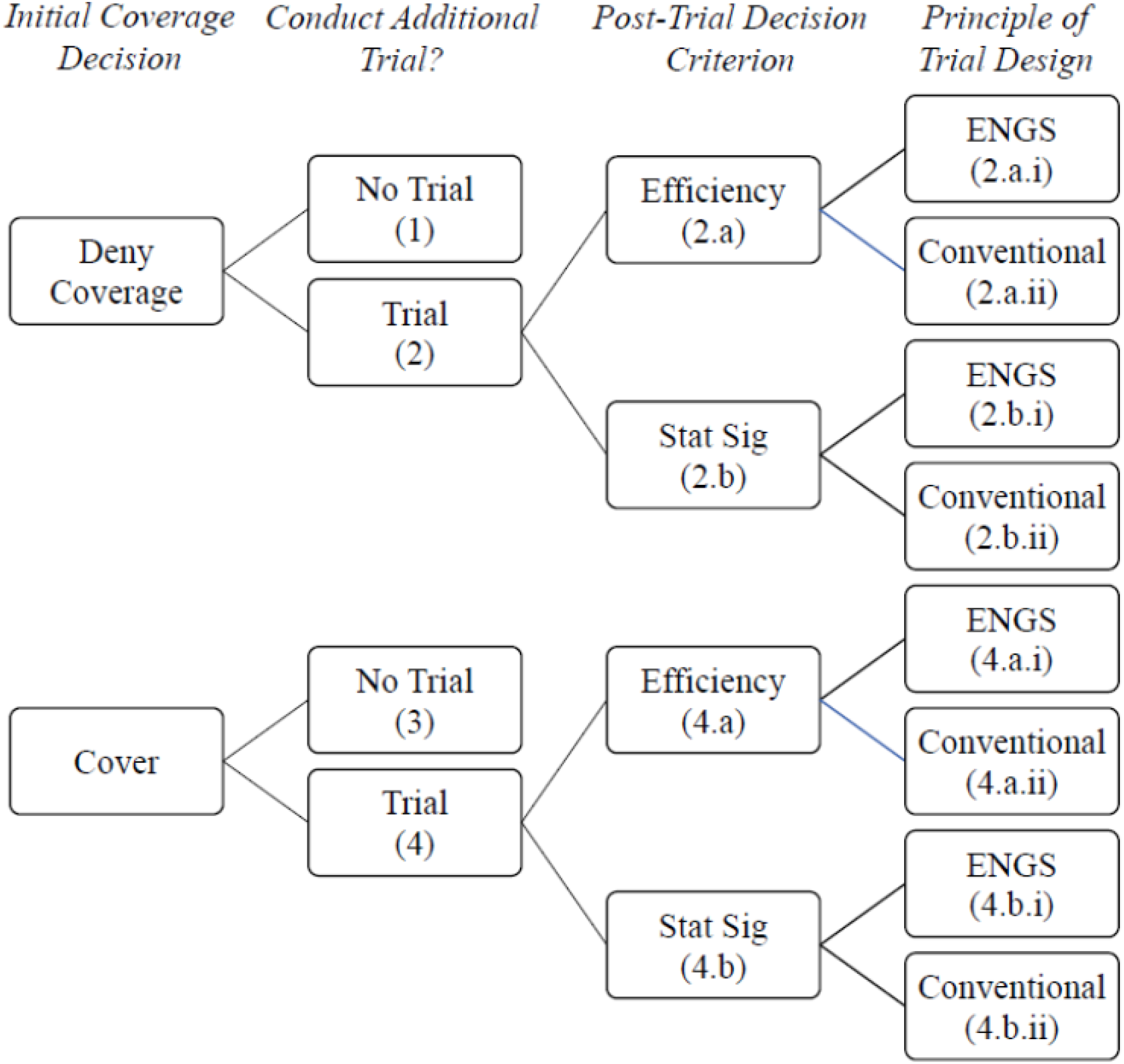
Schematic Representation of Decision Alternatives. Abbreviations: Stat Sig – Statistical Significance, ENGS – Expected Net Gain of Sampling Figure 1 depicts the decision alternatives considered in our analysis that would be available to a centralized decision maker. Because of the sequential nature of the decision-making process, alternatives are assigned a final number on the right side of the diagram when no further branches are possible. A trial refers to a randomized, placebo-controlled parallel trial that would be conducted to further clarify the efficacy of high-dose aducanumab for MCI and mild AD patients. The post-trial decision criterion refers to the principles used by the decision maker to interpret the results of a trial and update its coverage decision. Efficiency implies considering costs as well as health effects, with the specific criterion for post-trial coverage based on whether expected (population) INMB>0. Statistical Significance implies aducanumab would be covered after a trial if and only if there was a statistically significant (p<0.05) reduction in the hazard of disease progression among MCI patients. The principle of trial design refers to the approach used to select two components of trial design: sample size and follow-up length. ENGS implies the design with the largest positive ENGS (assuming one exists) is selected. Conventional implies an 18-month RCT is conducted with sample size set by statistical power calculations.

Initial debate focused on whether CMS should cover aducanumab for the general patient population or not, with CED suggested as an attractive compromise.^6,11^ However, given the pessimism surrounding aducanumab’s clinical efficacy^12-16^ and its high cost and that of RCTs^17^, it is reasonable to ask whether, given current knowledge, a further RCT is an efficient use of limited resources and thus whether CED was the optimal choice compared with available alternatives. The manufacturer has pushed ahead with a phase 4 confirmatory RCT - ‘ENVISION’ (ClinicalTrials.gov: NCT05310071) - with results expected by the end of 2026. However, it is unclear if the manufacturer’s interests align with those of society. And if not, what is the expected opportunity loss attributable to conducting a trial and, indirectly, to CED.

We expand on ICER’s results by performing a modeling-based expected value of sample information (EVSI)^18^ analysis. We quantify the decision-theoretic value of an RCT evaluating the clinical efficacy of aducanumab and determine what sized trial, if any, should be conducted, conditional on an initial coverage decision and CMS’s post-trial decision criterion. We also evaluate an RCT analogous to the ENVISION trial. Our results allow us to formally rank the four decision alternatives and quantify the opportunity costs at stake in this decision.

## Methods

We compare the 4 decision alternatives in terms of US population-level expected net monetary benefit (NMB) over the decision timeframe – the period during which the decision is relevant.^19^ Our analysis involved three main steps: (I) *cost-utility analysis (CUA)*: estimate the per-person incremental NMB (INMB) associated with high-dose aducanumab compared with care as usual; (II) *EVSI analysis*: estimate the per-person and population-level expected value and costs of a range of trials and identify optimal designs; and (III) *evaluating decision alternatives:* aggregate (I) to the population level and integrate with (II). Below we briefly describe our model and our approach to I-III. Full details are given in the Supplementary methods.

### Model

We re-implemented the decision-analytic model described in ICER’s report^6^ to characterize the clinical course of AD with and without aducanumab. In particular, we created a stochastic discrete-time cohort Markov model^20^ that uses similar structure, parameter point estimates, and costs as in the ICER model (see Figure 2 for our model structure). We independently assessed all major modeling decisions; for some parameters, e.g., caregiver quality of life, we used different values or made different modeling decisions than the ICER report. When, on the rare occasion, ICER did not report specific parameter values, we informed needed parameters from the literature. In general, we specified uncertainty distributions for parameters independently of ICER’s report due to incomplete information or to account for important parameter dependence. Full details of our model are given in Supplementary methods and eTables 1-4. The model is implemented in R version 4.1.1.^21^ It underwent extensive internal and cross (with the ICER model) validation to ensure that it is implemented correctly and that it behaves as expected.^22^ Cross-validation results are available upon request.

**Figure 2:**
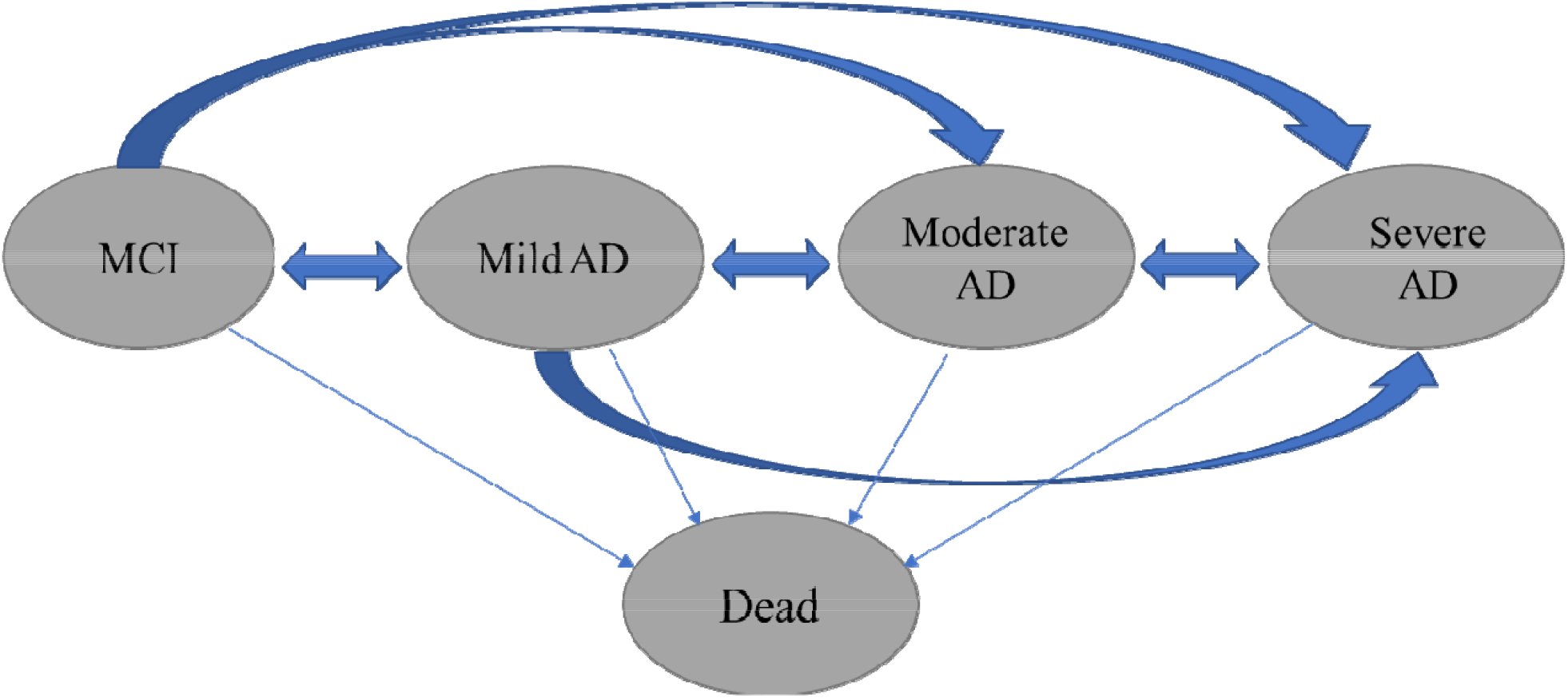
Model States and Transitions. Abbreviations: AD – Alzheimer’s Disease, MCI – Mild Cognitive Impairment The model relies on five basic types of states: MCI (due to AD), mild, moderate, and severe AD, and death. Separate states are used for combinations of setting (community-dwelling vs. long-term care), sex (male/female), and Aducanumab treatment-status/history (no-treatment, treatment, and discontinued treatment). For each AD severity state, within-cycle transitions are allowed to any more advanced AD state (e.g., MCI to moderate AD), the preceding less-severe state, (e.g., mild AD to MCI), and death. Mortality rates are higher than the general population and increase with AD severity.

The model simulates the time-varying distribution of an initial cohort of patients across five states – MCI, mild, moderate, and severe AD, and death - using 3-month cycles. We assume treated patients receive high-dose aducanumab (titrated to 10 mg/kg of bodyweight) while in the MCI or mild AD state. Its effect is implemented via a hazard ratio (HR) on the baseline hazard rate of progressing to any more advanced AD state, with a separate HR for MCI and mild AD. For our reference analysis, we conducted a Bayesian meta-analysis of the two trials to estimate the MCI-specific HR and used the meta-analytic predictive distribution to model uncertainty (‘current knowledge’). In the absence of data, we follow ICER in assuming a 50% reduction in the effectiveness of treatment for mild AD (vs. MCI) patients, and we model uncertainty for this parameter with a continuous uniform distribution over (0, 1). Following ICER^6^ and a manufacturer-sponsored analysis^23^, we also consider an optimistic scenario using only the EMERGE trial results and assuming equal efficacy for mild AD patients.

The outcomes of the model include life years (LYs), quality-adjusted life years (QALYs), and costs. We performed all analyses from both the societal and US healthcare perspective. For analyses undertaken from the US healthcare perspective, only patient QALYs are considered, but, for the societal perspective, the sum of caregiver and patient QALYs is used.^24^ For the cost of aducanumab, we use the recently reduced estimated wholesale acquisition price (WAP) - $28.2K (thousand) per year/per patient (original=$56K).^5^ Full details about costs are given in eTable 5. All costs are given in 2020 US dollars. All analyses were performed both undiscounted and using a 3%^25^ discount rate for costs and benefits.

### Per-Person CUA

We used a lifetime time horizon to estimate the per-person mean LYs, QALYs, and total costs for a mixed cohort of early-stage AD patients – MCI (55%) and mild AD (45%) patients in the community (92%) and a LTC setting (8%) – with and without treatment with high-dose aducanumab beginning at age 70 (52% female). To compare per-person mean incremental health gains and costs attributable to initiating treatment, we calculated (eEquation 1) the per-person mean INMB using willingness to pay (WTP) values (for a QALY) between $50K-$500K. However, in this analysis, and all others, we focus on WTP values between $50K-$200K, a range roughly in line with empirical and theoretical research.^26-28^ A positive (negative) INMB entails the expected additional benefits of treatment are greater (less) than the additional expected costs for a given WTP.

We propagated uncertainty in the input parameters through the model via forward Monte-Carlo to derive uncertainty in model outputs (often termed “probabilistic sensitivity analysis”^29^). For each outcome, we report the mean and 95% credible intervals based on the 0.025 and 0.975 quantiles of the uncertainty distribution. We also present both a cost-effectiveness acceptability curve (CEAC) and frontier (CEAF) across the range of considered WTP values.^30^ Finally, we investigated how the posterior mean per-person INMB would vary with the MCI-specific HR.

### EVSI Analysis

We used an EVSI framework to quantify the expected benefit, given current knowledge, of conducting further research on the clinical efficacy of aducanumab and, through comparison with expected trial-related costs, to evaluate alternative designs (including not doing a trial). An EVSI framework focuses on the decision-theoretic value of a trial: its ability to help improve decision making. Specifically, the expected benefit of a trial (eEquations 2-4) is the difference between the expected value of a (coverage) decision made after completing the trial and one made based on current knowledge, where the value of each decision is the total QALYs it produces net the opportunity cost of the resources it consumes, i.e., NMB. A trial potentially confers this type of value for each patient whose treatment decision might be impacted by the results, and, thus, the total benefit of a trial is the sum of the per-person benefit across the population over the decision timeframe (eEquation 5). However, as with health interventions and technology, expensive trials preclude using the same resources for other trials and thus the benefit of a trial must be weighed against its costs. The costs of a trial include the opportunity cost (eEquation 6) of treating a subset of participants with the non-standard treatment, e.g., half of participants are randomized to placebo and thereby forego the opportunity to undergo treatment, and direct research costs (eEquation 7).^18^ Finally, to integrate benefit and costs, we calculate the net difference - the “expected net gain from sampling” (ENGS).^31^ In an EVSI framework, the optimal trial design is that which maximizes the ENGS, or no trial (N=0) if no design has positive ENGS.

We evaluated a balanced, parallel-group and placebo-controlled RCT designed to reduce uncertainty about the treatment effectiveness HRs (MCI and mild AD). We considered variable follow-up lengths (12-36 months in 6-month increments) and total trial sample sizes (1K-10K, by increments of 100). For each combination, we estimated the per-person and population-level EVSI (using an established approximation method^32^ for the former), total trial costs, and ENGS. We conditioned all analyses on an initial coverage decision: don’t cover or cover.

The real-world (decision-theoretic) expected value of a trial implicitly depends on the decision criterion used by policymakers to interpret results and subsequently determine the (new) coverage policy. We considered two idealized post-trial decision criteria: (a) after incorporating the new evidence via Bayesian inference, are health benefits expected to outweigh costs (hereon termed “efficiency”) and (b) did the new trial demonstrate a statistically significant (p<0.05) reduction in the rate of AD progression (hereon “statistical significance”). The former is standard in an EVSI framework while the latter is meant to represent a scenario where only clinical benefit is considered (ignoring costs) and CMS utilizes a common criterion in the clinical community to demarcate a ‘real effect’ (statistical significance).

To aggregate per-person EVSI results to the population level, we assumed 50K patients would take aducanumab the first year it was covered.^5^ We further assumed that this number would double (to 100K patients) the second year, and remain constant in all subsequent years, due to increased availability. However, we repeated the analyses assuming only 50K and up to 150K patients would begin treatment in each subsequent year. We used a 15-year timeframe but assessed the impact of 10- and 20-year timeframe. The health benefits and costs of future patients were discounted at the same rate as the per-person analysis (0% or 3%). To assess the dependency of our results on population size, we conducted a one-way sensitivity analysis on the implied population size (based on decision timeframe and potential annual treatment rates). To predict the total expected direct research costs of a trial as a function of sample size and duration, we used an estimate of the unit cost per patient per visit to a research clinic ($3,934) which was taken from a costing analysis of recent RCTs conducted to evaluate a novel therapeutic agent.^33^ Trial opportunity costs were derived from the cost-utility results. Finally, we identified the optimal trial for each analysis by the criterion of maximum positive ENGS. However, we also calculated the ENGS for an ENVISION-like trial: 18 months follow up and a sample size determined by a statistical power analysis (Type I error=0.05; Type II error=0.1)

To partially assess the sensitivity of our EVSI results to the cost and efficacy of aducanumab, we repeated the above analyses under two hypothetical price-reduction scenarios - assuming the WAP is reduced to $14.1K (another 50% reduction) and $5.6K (10% of its original value) – and the optimistic efficacy scenario.

### Evaluating Decision Alternatives

For each decision alternative, we used the above results to calculate the expected INMB (reference=NoTrt/NoTrial) for the US population of patients who might initiate aducanumab over the decision time frame (eEquations 8-10). We quantified the expected loss (forgone population NMB) associated with selecting each alternative as the difference between its expected population INMB and that of the optimal choice. For one set of analyses, we assumed alternatives with trials (eEquations 9-10) would use the optimal (or least bad) trial (by the criterion of ENGS) for each considered WTP. However, we also repeated the analysis assuming an ENVISION-like trial would be performed

## Results

### Per-Person CUA

Cost-utility results are given in the Appendix (eTable 6-8, eFigure 1-4). For the reference analysis, the expected per-person INMB of high-dose aducanumab compared with usual care ranged from -$93K to -$32K (additional costs outweigh benefits) depending on WTP. Parameter uncertainty does not translate into significant decision uncertainty. Even at a WTP of $200K, we predict <5% chance treatment is optimal. This result is robust to the considered alternative scenarios – including the optimistic scenario - except for WAP=$5.6K where treatment is optimal for WTP≥$200K.

### EVSI Analysis

In all analyses, a large population of AD patients - 1.45M (million) undiscounted people in the reference analysis - was assumed to be affected by an initial coverage decision (300K pre-trial) and possible trial results (1.15M post-trial), and, so, even a small per-person EVSI was amplified at the population level. Detailed expected trial costs are given in eTable 9-10: with (reference-case) direct research costs ranging from $19M (million) for a smaller 12-month trial (N=1,000) to $480M for a large 36-month trial (N=10,000). Examples of how trial costs and population EVSI are combined and used to select the trial with the maximum ENGS are given in eFigures 5-8, and power analysis results are presented in eFigure 9. Optimal trial design results from the reference analysis (societal perspective) are given in Table 1, and a summary of the impact of alternative modeling scenarios on these results is displayed in Table 2. Results differ notably depending on CMS’s initial coverage decision. Below we address each scenario in turn.

**Table 1:**
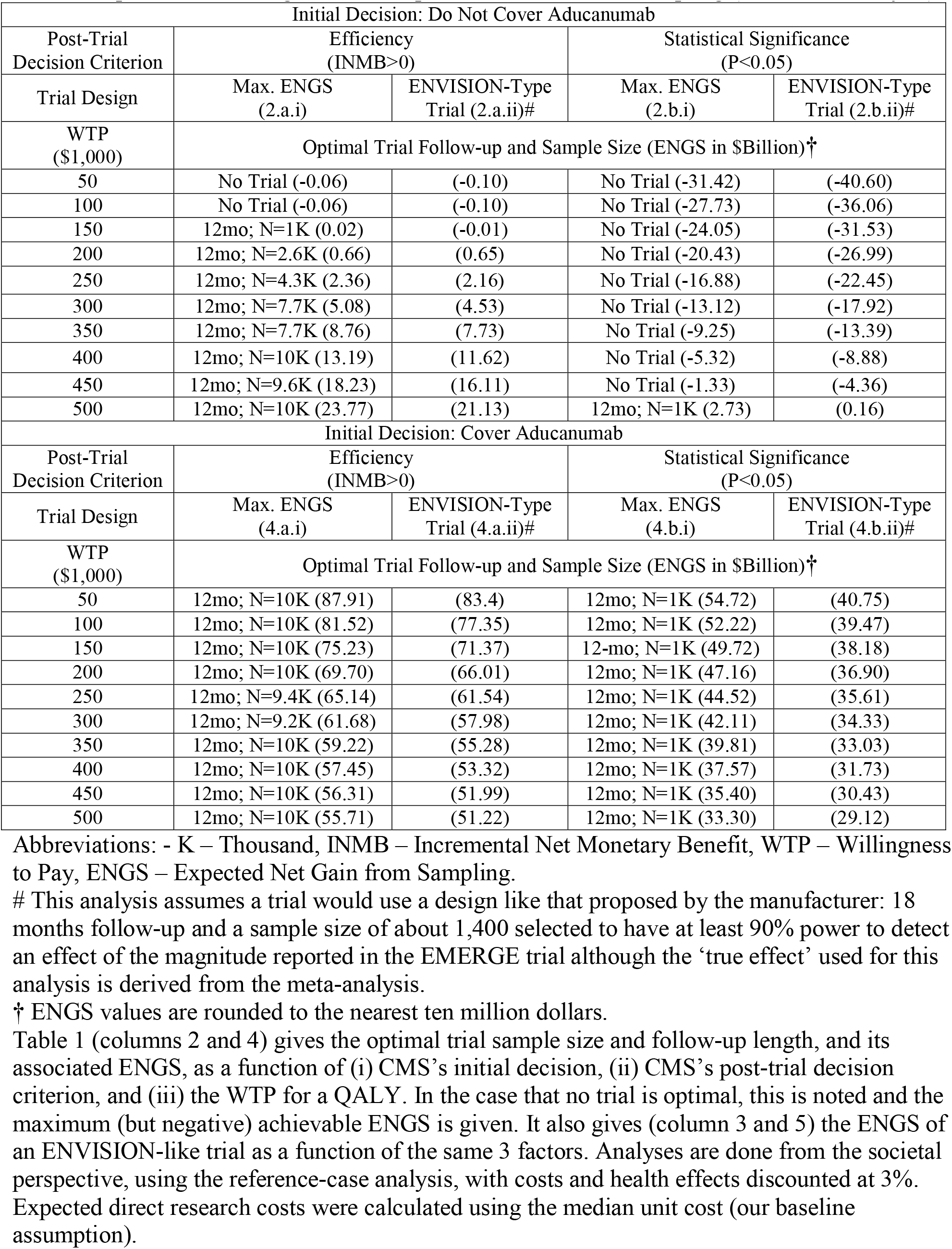
Optimal Trial Design and the Expected Net Gain from Sampling (Reference Analysis)

| Initial Decision: Do Not Cover Aducanumab |  |  |  |  |
| --- | --- | --- | --- | --- |
| Post-Trial Decision Criterion | Efficiency (INMB>0) |  | Statistical Significance (P<0.05) |  |
| Trial Design | Max. ENGS (2.a.i) | ENVISION-Type Trial (2.a.ii)# | Max. ENGS (2.b.i) | ENVISION-Type Trial (2.b.ii)# |
| WTP (\$1,000) | Optimal Trial Follow-up and Sample Size (ENGS in \$Billion)† | | | |
| 50 | No Trial (-0.06) | (-0.10) | No Trial (-31.42) | (-40.60) |
| 100 | No Trial (-0.06) | (-0.10) | No Trial (-27.73) | (-36.06) |
| 150 | 12mo; N=1K (0.02) | (-0.01) | No Trial (-24.05) | (-31.53) |
| 200 | 12mo; N=2.6K (0.66) | (0.65) | No Trial (-20.43) | (-26.99) |
| 250 | 12mo; N=4.3K (2.36) | (2.16) | No Trial (-16.88) | (-22.45) |
| 300 | 12mo; N=7.7K (5.08) | (4.53) | No Trial (-13.12) | (-17.92) |
| 350 | 12mo; N=7.7K (8.76) | (7.73) | No Trial (-9.25) | (-13.39) |
| 400 | 12mo; N=10K (13.19) | (11.62) | No Trial (-5.32) | (-8.88) |
| 450 | 12mo; N=9.6K (18.23) | (16.11) | No Trial (-1.33) | (-4.36) |
| 500 | 12mo; N=10K (23.77) | (21.13) | 12mo; N=1K (2.73) | (0.16) |
| Initial Decision: Cover Aducanumab |  |  |  |  |
| Post-Trial Decision Criterion | Efficiency (INMB>0) |  | Statistical Significance (P<0.05) |  |
| Trial Design | Max. ENGS (4.a.i) | ENVISION-Type Trial (4.a.ii)# | Max. ENGS (4.b.i) | ENVISION-Type Trial (4.b.ii)# |
| WTP (\$1,000) | Optimal Trial Follow-up and Sample Size (ENGS in \$Billion)† | | | |
| 50 | 12mo; N=10K (87.91) | (83.4) | 12mo; N=1K (54.72) | (40.75) |
| 100 | 12mo; N=10K (81.52) | (77.35) | 12mo; N=1K (52.22) | (39.47) |
| 150 | 12mo; N=10K (75.23) | (71.37) | 12mo; N=1K (49.72) | (38.18) |
| 200 | 12mo; N=10K (69.70) | (66.01) | 12mo; N=1K (47.16) | (36.90) |
| 250 | 12mo; N=9.4K (65.14) | (61.54) | 12mo; N=1K (44.52) | (35.61) |
| 300 | 12mo; N=9.2K (61.68) | (57.98) | 12mo; N=1K (42.11) | (34.33) |
| 350 | 12mo; N=10K (59.22) | (55.28) | 12mo; N=1K (39.81) | (33.03) |
| 400 | 12mo; N=10K (57.45) | (53.32) | 12mo; N=1K (37.57) | (31.73) |
| 450 | 12mo; N=10K (56.31) | (51.99) | 12mo; N=1K (35.40) | (30.43) |
| 500 | 12mo; N=10K (55.71) | (51.22) | 12mo; N=1K (33.30) | (29.12) |

Abbreviations: - K – Thousand, INMB – Incremental Net Monetary Benefit, WTP – Willingness to Pay, ENGS – Expected Net Gain from Sampling.

**Table 2:** Impact of Scenarios on Optimal Trial Design and the ENGS of a Manufacturer Trial.

| Modeling Scenario or Assumption | Optimal Trial Design (ENGS# in \$Billion) given CMS's Initial Decision Not to Cover Aducanumab and Assuming a Post-Trial Decision Criterion of Efficiency (INMB>0) ¶ | | | | |
| --- | --- | --- | --- | --- | --- |
| | WTP (\$1,000) | | | | |
|  | 50 | 100 | 150 | 200 | 250 |
| Reference Analysis | No Trial (-0.06) | No Trial (-0.06) | 12mo; N=1K (0.02) | 12mo; N=2.6K (0.66) | 12mo; N=4.3K (2.36) |
| Optimistic Analysis | No Trial (-0.08) | No Trial (-0.06) | No Trial (-0.05) | No Trial (-0.04) | 12mo; N=10K (2.49) |
| WAP=\$14,100 | No Trial (-0.04) | 18mo; N=1K (0.33) | 12mo; N=4.3K (2.38) | 12mo; N=7.8K (6.18) | 12mo; N=9.4K (11.21) |
| WAP=\$5,600 | 12mo; N=2.2K (0.40) | 12mo; N=5.5K (3.88) | 12mo; N=7.8K (9.64) | 12mo; N=8.8K (16.51) | 12mo; N=10K (23.86) |
| 10-Yr Timeframe (Reference) | No Trial (-0.06) | No Trial (-0.06) | No Trial (-0.01) | 12mo; N=1K (0.36) | 12mo; N=2.8K (1.35) |
| 20-Yr Timeframe (Reference) | No Trial (-0.06) | No Trial (-0.06) | 12mo; N=1K (0.04) | 18mo; N=2.3K (0.94) | 12mo; N=4.4K (3.23) |
| 50K Treated/Yr (Reference) | No Trial (-0.06) | No Trial (-0.06) | No Trial (-0.02) | 12mo; N=1K (0.31) | 12mo; N=2.7K (1.15) |
| 150K Treated/Yr (Reference) | No Trial (-0.06) | No Trial (-0.06) | 12mo; N=1K (0.05) | 12mo; N=3.2K (1.05) | 12mo; N=4.4K (3.58) |
| Modeling Scenario or Assumption | ENGS (\$Billion) for Manufacturer's Trial given Initial Decision Not to Cover Aducanumab and Assuming a Post-Trial Criterion of Efficiency or (Statistical Significance; P<0.05) ¶ | | | | |
| | WTP (\$1,000) | | | | |
|  | 50 | 100 | 150 | 200 | 250 |
| Reference Analysis | -0.10 (-40.60) | -0.10 (-36.06) | -0.01 (-31.53) | 0.65 (-26.99) | 2.16 (-22.45) |
| Optimistic Analysis | -0.12 (-80.19) | -0.10 (-63.53) | -0.09 (-46.88) | -0.05 (-30.24) | 1.33 (-13.62) |
| WAP=\$14,100 | -0.07 (-19.47) | 0.33 (-14.93) | 2.16 (-10.40) | 5.49 (-5.88) | 9.99 (-1.36) |
| WAP=\$5,600 | 0.39 (-6.73) | 3.49 (-2.21) | 8.71 (2.33) | 14.99 (6.88) | 21.76 (11.41) |
| 10-Yr Timeframe † | -0.10 (-23.82) | -0.10 (-21.16) | -0.04 (-18.51) | 0.35 (-15.85) | 1.23 (-13.19) |
| 20-Yr Timeframe † | -0.10 (-55.06) | -0.10 (-48.91) | 0.02 (-42.76) | 0.92 (-36.61) | 2.96 (-30.45) |
| 50K Treated/Yr † | -0.10 (-21.43) | -0.10 (-19.04) | -0.05 (-16.65) | 0.30 (-14.26) | 1.10 (-11.86) |
| 150K Treated/Yr † | -0.10 (-59.76) | -0.10 (-53.09) | 0.03 (-46.41) | 1.00 (-39.73) | 3.32 (-33.05) |

Abbreviations: - CMS – Centers for Medicare and Medicaid Services, WTP – Willingness to Pay, N – Sample Size, K – Thousands, ENGS – Expected Net Gain from Sampling, WAP – Wholesale Acquisition Price, Yr – Year
¶If no trial is optimal (there is no design with ENGS>0), the ENGS of the least-bad trial is given. Table 2 shows (top) how optimal trial design and the associated ENGS would vary under different modeling scenarios conditional on CMS's actual decision to not cover aducanumab and assuming CMS would rely on an efficiency-based post-trial decision criterion. For each scenario, the optimal trial design and associated ENGS are given for a range of WTP values. In each case, the optimal trial design is that with the largest positive ENGS or no trial if all ENGS<0. Table 2 also shows (bottom) how the ENGS associated with an ENVISION-like trial (18-month follow up and roughly 1,400 enrollees) would vary under the same modeling scenarios, conditional on CMS's no-coverage decision and assuming it would rely on efficiency or statistical significance (given in parentheses) for a post-trial decision criterion.
¶ Analyses are done from the societal perspective with costs and health effects discounted at 3%. Expected direct research costs were calculated using the median unit cost (our baseline assumption).
†All other analysis parameters are from the Reference analysis.

### Deny Coverage: Trial vs. No Trial

Given the actual initial decision to deny coverage, the net value of conducting an RCT and its optimal sample size depend on CMS’s post-trial decision criterion and, to a lesser extent, the WTP for a QALY. For WTP≤$100K and assuming a post-trial decision criterion of efficiency, none of the considered designs confer decision-theoretic value (EVSI=0), making no trial clearly optimal. At WTP=$150K, per-person EVSI is small (≈$80), and the smallest considered 12-month trial (N=1,000) optimal. However, ENGS<0 for (trial) N≥1,400, and the optimality of a trial is sensitive to the population size and treatment-efficacy uncertainty distribution. For an undiscounted population<1.2M (average of 80K/year for 15 years), no trial is optimal for WTP≤$150K. Moreover, under the optimistic scenario, EVSI=0 for WTP≤$150K. For WTP≥$200K, an increasingly large trial would be optimal (per-person EVSI≈$900) in the reference analysis for population>120K (8K/year) although not until WTP=$250K in the optimistic scenario. With a reduced drug cost WAP=$14.1K ($5.6K), per-person EVSI>0 for WTP≥$100K (WTP≥$50K) and an order of magnitude larger, and significantly larger trials are optimal over the same WTP ranges.

If CMS made its post-trial decision based on statistical significance, even small trials would carry substantial negative value (per-person EVSI≤-$25K for WTP≤$200K) and no trial would be optimal for WTP ≤$450K. Per-person loss nearly doubles under the optimistic scenario for WTP ≤$100K. On the population level, conducting an ENVISION-like trial would entail an ENGS between -$25B (billion) and -$40B for WTP≤$200K. This is robust to considered scenarios except for a large WAP reduction. An ENVISION-type trial would have positive ENGS beginning at WTP=$150K for WAP=$5.6K.

### Offer Coverage: Trial vs. No Trial

If CMS had instead chosen to broadly cover high-dose aducanumab, the ENGS of an RCT would be billions of dollars (per-person EVSI between $30K-$100K). See Appendix for further results.

### Evaluating Decision Alternatives

Table 3 and eTable 11 give the ranking of the four decision alternatives (reference analysis, societal perspective) and the expected opportunity loss of suboptimal decisions conditional on CMS’s post-trial decision criterion, the principle of trial design, and WTP. Table 4 and eTable 12 outline how these results are influenced by alternative modeling scenarios. We highlight 4 key results. First, adopting treatment would have carried a substantial opportunity cost with (>$16B) or without (>$86B) a trial. Second, for WTP≤$100K or a post-trial decision criterion of statistical significance, NoTrt/NoTrial is optimal barring a large (≥50%) price reduction. Third, at WTP=$150K (efficiency), NoTrt/Trial is optimal, but not robustly so. However, although with the ENVISION trial NoTrt/NoTrial is optimal, the expected loss of doing the trial is small ($10M) relative to population size. For WTP≥$200K (efficiency), NoTrt/Trial is (more robustly) optimal and the loss associated with NoTrt/NoTrial is large (≥$650M). Moreover, for WAP=$5.6K, NoTrt/Trial is optimal for WTP≤$150K, and Trt/Trial is optimal for WTP≥$200K.

**Table 3:** Ranking of Decision Alternatives and Expected Opportunity Loss.

| Post-Trial Decision Criterion | Efficiency (INMB>0) |  | Statistical Significance (P<0.05) |  |
| --- | --- | --- | --- | --- |
| Trial Design | Max. ENGS (2.a.i/4.a.i)# | ENVISION-Type Trial (2.a.ii/4.a.ii)† | Max. ENGS (2.b.i/4.b.i)# | ENVISION-Type Trial (2.b.ii/4.b.ii)† |
| WTP†† (\$1,000) | Ranking (Expected Opportunity Loss of selecting a suboptimal decision given as forgone \$Billions of NMB*) of Each Decision Alternative.¶ | | | |
| 50 | 1.NoTrt/NoTrial<br>2.NoTrt/Trial (0.06)<br>3.Trt/Trial (21.83)<br>4.Trt/NoTrial (110.20) | 1.NoTrt/NoTrial<br>2.NoTrt/Trial (0.10)<br>3.Trt/Trial (26.73)<br>4.Trt/NoTrial (110.20) | 1.NoTrt/NoTrial<br>2.NoTrt/Trial (31.42)<br>3.Trt/Trial (55.43)<br>4.Trt/NoTrial (110.20) | 1.NoTrt/NoTrial<br>2.NoTrt/Trial (40.59)<br>3.Trt/Trial (69.38)<br>4.Trt/NoTrial (110.20) |
| 100 | 1.NoTrt/NoTrial<br>2.NoTrt/Trial (0.06)<br>3.Trt/Trial (20.26)<br>4.Trt/NoTrial (102.20) | 1.NoTrt/NoTrial<br>2.NoTrt/Trial (0.10)<br>3.Trt/Trial (24.79)<br>4.Trt/NoTrial (102.20) | 1.NoTrt/NoTrial<br>2.NoTrt/Trial (27.73)<br>3.Trt/Trial (49.93)<br>4.Trt/NoTrial (102.20) | 1.NoTrt/NoTrial (0.00)<br>2.NoTrt/Trial (36.06)<br>3.Trt/Trial (62.67)<br>4.Trt/NoTrial (102.20) |
| 150 | 1.NoTrt/Trial<br>2.NoTrt/NoTrial (0.02)<br>3.Trt/Trial (18.60)<br>4.Trt/NoTrial (94.40) | 1.NoTrt/NoTrial<br>2.NoTrt/Trial (0.01)<br>3.Trt/Trial (22.77)<br>4.Trt/NoTrial (94.40) | 1.NoTrt/NoTrial<br>2.NoTrt/Trial (24.05)<br>3.Trt/Trial (44.44)<br>4.Trt/NoTrial (94.20) | 1.NoTrt/NoTrial<br>2.NoTrt/Trial (31.53)<br>3.Trt/Trial (55.97)<br>4.Trt/NoTrial (94.20) |
| 200 | 1.NoTrt/Trial<br>2.NoTrt/NoTrial (0.66)<br>3.Trt/Trial (16.81)<br>4.Trt/NoTrial (86.86) | 1.NoTrt/Trial<br>2.NoTrt/NoTrial (0.65)<br>3.Trt/Trial (20.79)<br>4.Trt/NoTrial (86.86) | 1.NoTrt/NoTrial<br>2.NoTrt/Trial (20.43)<br>3.Trt/Trial (39.01)<br>4.Trt/NoTrial (86.20) | 1.NoTrt/NoTrial<br>2.NoTrt/Trial (26.99)<br>3.Trt/Trial (49.26)<br>4.Trt/NoTrial (86.20) |
| 250 | 1.NoTrt/Trial<br>2.NoTrt/NoTrial (2.36)<br>3.Trt/Trial (15.11)<br>4.Trt/NoTrial (80.56) | 1.NoTrt/Trial<br>2.NoTrt/NoTrial (2.16)<br>3.Trt/Trial (18.78)<br>4.Trt/NoTrial (80.36) | 1.NoTrt/NoTrial<br>2.NoTrt/Trial (16.88)<br>3.Trt/Trial (33.65)<br>4.Trt/NoTrial (78.20) | 1.NoTrt/NoTrial<br>2.NoTrt/Trial (22.45)<br>3.Trt/Trial (42.54)<br>4.Trt/NoTrial (78.20) |
| 300 | 1.NoTrt/Trial<br>2.NoTrt/NoTrial (5.08)<br>3.Trt/Trial (13.34)<br>4.Trt/NoTrial (75.29) | 1.NoTrt/Trial<br>2.NoTrt/NoTrial (4.53)<br>3.Trt/Trial (16.72)<br>4.Trt/NoTrial (74.74) | 1.NoTrt/NoTrial<br>2.NoTrt/Trial (13.12)<br>3.Trt/Trial (28.07)<br>3.Trt/NoTrial (70.20) | 1.NoTrt/NoTrial<br>2.NoTrt/Trial (17.92)<br>3.Trt/Trial (35.84)<br>4.Trt/NoTrial (70.21) |

Abbreviations: NMB – Net Monetary Benefit, INMB – Incremental NMB, Max – Maximum, ENGS – Expected Net Gain from Sampling, WTP – Willingness to Pay, NoTrt – Do Not Cover Aducanumab, Trt – Cover Aducanumab, NoTrial – Do Not Perform Trial, Trial – Perform Trial.
#In columns 2/4, decision alternatives including a trial are assumed to include the optimally designed trial, where optimality is defined in terms of ENGS and dependent on the WTP for a QALY and CMS's initial coverage decision and assumed post-trial decision criterion. In cases where no trial is optimal, i.e., no trial has ENGS>0, the least bad trial is used (smallest loss).
†In these analysis, any decision alternative that includes a trial, e.g., Trt/Trial, is assumed to include a trial designed to have at least 90% power to detect the effect size reported in the EMERGE trial given 18 months of follow-up (N≈1,400). Such a trial is comparable to the upcoming phase-4 trial ('ENVISION') planned by the manufacturer.
\*Expected opportunity losses represent forgone NMB (rounded to the nearest \$10,000,000) associated with selecting each option instead of the best.
¶ Analyses are done from the societal perspective, using the reference-case analysis, with costs and health effects discounted at 3%. Expected direct research costs were calculated using the median unit cost (our baseline assumption).

**Table 4:** Impact of Modeling Scenarios on the Decision Alternative Rankings.

| Modeling Scenario or Assumption | Ranking (Expected Opportunity Loss of selecting a suboptimal decision given as forgone \$Billions of NMB*) of Each Decision Alternative Assuming Optimal Trials (max ENGS)# and a Post-Trial Decision Criterion of Efficiency (INMB>0)¶ | | | |
| --- | --- | --- | --- | --- |
| | WTP (\$1,000) | | | |
|  | 50 | 100 | 150 | 200 |
| Reference Analysis | 1.NoTrt/NoTrial<br>2.NoTrt/Trial (0.06)<br>3.Trt/Trial (21.83)<br>4.Trt/NoTrial (110.20) | 1.NoTrt/NoTrial<br>2.NoTrt/Trial (0.06)<br>3.Trt/Trial (20.26)<br>4.Trt/NoTrial (102.20) | 1.NoTrt/Trial<br>2.NoTrt/NoTrial (0.02)<br>3.Trt/Trial (18.60)<br>4.Trt/NoTrial (94.22) | 1.NoTrt/Trial<br>2.NoTrt/NoTrial (0.66)<br>3.Trt/Trial (16.81)<br>4.Trt/NoTrial (86.86) |
| Optimistic Analysis | 1.NoTrt/NoTrial<br>2.NoTrt/Trial (0.08)<br>3.Trt/Trial (27.09)<br>4.Trt/NoTrial (136.97) | 1.NoTrt/NoTrial<br>2.NoTrt/Trial (0.06)<br>3.Trt/Trial (21.90)<br>4.Trt/NoTrial (110.57) | *1.NoTrt/NoTrial* §<br>*2.NoTrt/Trial (0.05)*<br>3.Trt/Trial (16.72)<br>4.Trt/NoTrial (84.18) | *1.NoTrt/NoTrial* §<br>*2.NoTrt/Trial (0.04)*<br>2.Trt/Trial (11.44)<br>3.Trt/NoTrial (57.78) |
| WAP of \$14,100 | 1.NoTrt/NoTrial<br>2.NoTrt/Trial (0.04)<br>3.Trt/Trial (11.09)<br>4.Trt/NoTrial (55.48) | *1.NoTrt/Trial* §<br>*2.NoTrt/NoTrial (0.33)*<br>3.Trt/Trial (9.34)<br>4.Trt/NoTrial (47.82) | 1.NoTrt/Trial<br>2.NoTrt/NoTrial (2.38)<br>3.Trt/Trial (7.57)<br>4.Trt/NoTrial (41.87) | *1.NoTrt/Trial* §<br>*2.Trt/Trial (5.73)*<br>*3.NoTrt/NoTrial (6.18)*<br>4.Trt/NoTrial (37.67) |
| WAP of \$5,600 | *1.NoTrt/Trial* §<br>*2.NoTrt/NoTrial (0.40)*<br>3.Trt/Trial (4.48)<br>4.Trt/NoTrial (22.90) | *1.NoTrt/Trial* §<br>*2.Trt/Trial (2.61)*<br>*3.NoTrt/NoTrial (3.88)*<br>4.Trt/NoTrial (18.39) | *1.NoTrt/Trial* §<br>*2.Trt/Trial (0.76)*<br>*3.NoTrt/NoTrial (9.64)*<br>4.Trt/NoTrial (16.15) | *1.Trt/Trial* §<br>*2.NoTrt/Trial (1.14)*<br>*3.Trt/NoTrial (16.16)*<br>*4.NoTrt/NoTrial (17.66)* |
| Modeling Scenario or Assumption | Ranking (Expected Opportunity Loss of selecting a suboptimal decision given as forgone \$Billions of NMB*) of Each Decision Alternative Assuming an ENVISION-like Trial† and a Post-Trial Decision Criterion of Statistical Significance (P<0.05)¶ | | | |
| | WTP (\$1,000) | | | |
|  | 50 | 100 | 150 | 200 |
| Reference Analysis | 1.NoTrt/NoTrial<br>2.NoTrt/Trial (40.59)<br>3.Trt/Trial (69.38)<br>4.Trt/NoTrial (110.20) | 1.NoTrt/NoTrial<br>2.NoTrt/Trial (36.02)<br>3.Trt/Trial (62.67)<br>4.Trt/NoTrial (102.20) | 1.NoTrt/NoTrial<br>2.NoTrt/Trial (31.53)<br>3.Trt/Trial (55.97)<br>4.Trt/NoTrial (94.20) | 1.NoTrt/NoTrial<br>2.NoTrt/Trial (26.99)<br>3.Trt/Trial (49.26)<br>4.Trt/NoTrial (86.20) |
| Optimistic Analysis | 1.NoTrt/NoTrial<br>2.NoTrt/Trial (80.19)<br>3.Trt/Trial: (117.55)<br>4.Trt/NoTrial: (136.97) | 1.NoTrt/NoTrial<br>2.NoTrt/Trial (63.53)<br>3.Trt/Trial (93.63)<br>4.Trt/NoTrial (110.57) | 1.NoTrt/NoTrial<br>2.NoTrt/Trial (46.88)<br>3.Trt/Trial (69.71)<br>4.Trt/NoTrial (84.18) | 1.NoTrt/NoTrial<br>2.NoTrt/Trial (30.24)<br>3.Trt/Trial (45.81)<br>4.Trt/NoTrial (57.78) |
| WAP of \$14,100 | 1.NoTrt/NoTrial<br>2.NoTrt/Trial (19.47)<br>3.Trt/Trial (33.91)<br>4.Trt/NoTrial (55.48) | 1.NoTrt/NoTrial<br>2.NoTrt/Trial (14.93)<br>3.Trt/Trial (27.20)<br>4.Trt/NoTrial (47.49) | 1.NoTrt/NoTrial<br>2.NoTrt/Trial (10.40)<br>3.Trt/Trial (20.49)<br>4.Trt/NoTrial (39.49) | 1.NoTrt/NoTrial<br>2.NoTrt/Trial (5.88)<br>3.Trt/Trial (13.80)<br>4.Trt/NoTrial (31.49) |
| WAP of \$5,600 | 1.NoTrt/NoTrial<br>2.NoTrt/Trial (6.73)<br>3.Trt/Trial (12.53)<br>4.Trt/NoTrial (22.50) | 1.NoTrt/NoTrial<br>2.NoTrt/Trial (2.21)<br>3.Trt/Trial (5.83)<br>4.Trt/NoTrial (14.50) | *1.NoTrt/Trial* §<br>*2.Trt/Trial (1.45)*<br>*2.NoTrt/NoTrial (2.33)*<br>4.Trt/NoTrial (8.84) | *1.Trt/Trial* §<br>2.NoTrt/Trial (0.73)<br>*2.NoTrt/NoTrial (6.11)*<br>4.Trt/NoTrial (7.61) |

Abbreviations: INMB – Incremental Net Monetary Benefit, ENGS – Expected Net Gain from Sampling, WTP – Willingness to Pay, NoTrt – Do Not Cover, Trt – Cover Aducanumab, NoTrial – Do Not Perform Trial, Trial – Perform Trial, WAP – Wholesale Acquisition Price \*Values are rounded to the nearest billion dollars.
#In cases where no trial is optimal (no non-negative ENGS), the least bad trial is used.
†An 18-month follow up with a total sample size of roughly 1,400.
¶ Analyses are done from the societal perspective, using the reference-case analysis, with costs and health effects discounted at 3%. Expected direct research costs were calculated using the median unit cost (our baseline assumption).
§ Indicates the ranking differs from the reference case; relevant alternatives are italicized.

## Discussion

In this paper, we used a decision-theoretic framework to evaluate CMS’s national coverage policy for aducanumab (CED) by conceptualizing it as one (NoTrt/Trial) of four possible decision alternatives available to a centralized decision maker. To evaluate a trial, we quantified the degree to which it was expected to help a decision maker more efficiently allocate scarce healthcare-directed resources relative to its expected costs. However, our approach captured only a subset of this value - we ignored the value of reduced uncertainty concerning adverse events and focused only on the US jurisdiction - and excluded other important benefits of a trial, e.g., to guide future R&D decisions. With this limitation in mind, our results support the following conclusions, albeit with caveats. First, CMS wisely avoided the worst expected outcomes by deciding not to cover aducanumab for the general population. Second, for WTP≤$100K, a trial, and thus CED, is not expected to improve decision making and thus is not justified on decision-theoretic grounds given CMS’s initial coverage decision, current evidence, and the manufacturer-determined price. Third, for WTP=$150K and assuming an efficiency criterion, CED (with the ENVISION trial) is reasonable given the broader benefits of research and the small opportunity loss (relative to population size) for suboptimal trial design. Finally, at WTP≥$200K and efficiency, CED is optimal and justified on decision-theoretic grounds alone, and not doing a trial would amount to a costly missed opportunity to improve decision making by reducing uncertainty.

In our analyses, we used a stylized model of the decision-making process and considered two idealized post-trial decision criteria: efficiency (INMB>0) following Bayesian evidence updating and statistical significance (p<0.05). Conditioning on CMS’s initial decision to deny coverage, the expected (net) decision-theoretic value of a trial varied dramatically between the two criteria – opposite signs and 2x orders of magnitude. This is because, to justify the high costs of treatment on efficiency grounds, the estimated treatment effect would have to be very large and thus would necessarily fall in the tail of the uncertainty distribution. However, only a moderate-sized (and much more likely) true effect like that observed in the EMERGE trial (HR=0.69) would be needed to attain high statistical power. Thus, if CMS relied exclusively on the criterion of statistical significance, in most of the scenarios where a trial detected a benefit and lead to a coverage reversal, the true effect would not be sufficiently large to justify the high drug cost and thus the trial would precipitate a loss in NMB at the population level. Our conclusions therefore depend on which, if either, criterion is applicable to the real-world decision-making process.

By statute, CMS must cover applicable treatments deemed ‘reasonable and necessary’ and cannot explicitly rely on cost-effectiveness analyses. Thus, ostensibly, CMS would have to consider only the clinical impact of aducanumab in making it post-trial decision. Our efficiency scenario may still be informative if (i) CMS informally considers costs relative to benefit/harm profile under the umbrella of ‘reasonable’ or (ii) CMS looks for a large minimal clinically important difference to meet the standard of ‘necessary’. Significant uncertainty surrounding the clinical efficacy of aducanumab, in conjunction with nontrivial harms, surely provided cover for CMS policymakers to deny coverage without reference to the cost or budget impact of the drug. However, policymakers would presumably face significant pressure to amend their decision if a future trial reported an EMERGE-sized clinical benefit. Although justified from a efficiency standpoint, a decision to again deny coverage would likely be controversial and highly contested as, even in the face of current equivocal evidence, the manufacturer, patient-advocacy groups, and members of congress pressured CMS to amend their initial decision.^34^ Unfortunately, our results demonstrate that, at the current price, population-wide adoption of aducanumab to treat MCI and mild AD would entail a large expected opportunity loss (tens of $Billions) even if the EMERGE trial’s estimate was valid. Consequently, while it is needlessly speculative to predict the behavior of CMS, our results raise the possibility that even an ostensibly successful further trial (one that would surely be celebrated by the manufacturer and patient groups) could contribute to a large societal net loss if policymakers are not legally or politically free to incorporate the costs of treatment into their post-trial decision making process. Moreover, by incentivizing future research, CMS’s CED policy could directly contribute to this unfortunate outcome.

Our results also demonstrate how the high WAP set by the manufacture with exclusive market rights, in conjunction with current although fluid restrictions on CMS’s ability to leverage its monopsony market power to bargain down the price of new biologic therapies, undermines the economics of future research. For example, for WAP=$5.6K, a trial would be optimal for WTP=$150K regardless of CMS’s post-trial decision criterion and for all considered WTP if an efficiency criterion was used. This is because, at such a reduced price, a smaller, and thus more likely, effect size is required to achieve INMB>0. Treatment would be optimal at WTP=$100K for HR≤0.74 (vs. HR≤0.11 for WAP=$28.2K), at WTP=$150K for HR≤0.81 (vs. HR≤0.37), and at WTP=$200K for an effect size roughly equivalent to the meta-analytic point estimate, i.e., HR≤0.85 (vs. HR≤0.51).

The validity of our conclusions also depends on the validity of our meta-analysis of the EMERGE and ENGAGE MCI-specific HR estimates and of our reliance on it to characterize current knowledge about the clinical efficacy of high-dose aducanumab. In our analyses, we take the results of both trials at face value, and we ignore prior evidence. However, legitimate concerns have been raised about the internal validity of the trial results put forth by the manufacturer due to the partial (trial participants who did not complete the trial were excluded) and post-hoc nature of the analyses.^14^ Moreover, the manufacturer did not report a HR for the ENGAGE trial and so we relied on an estimate that ICER backed out from available data.

Beyond internal validity concerns, reasonable minds could disagree with our decision to use an uninformative prior distribution in the Bayesian meta-analysis. Two recent meta-analyses^15,16^ based on >25 RCTs of treatments targeting accumulated Aβ plaques have cast doubt on the underlying theory behind aducanumab – the amyloid cascade hypothesis. While these studies synthesize evidence across many different therapeutics, perhaps questionably, it certainly would be a defensible approach to at least consider the impact of formally incorporating this prior pessimistic evidence into the meta-analysis via an informative prior. In such a scenario, our results would likely portray the net value of a future trial (and thus CED) less favorably and thus would potentially corroborate the arguments made by authors questioning the ethics of future trials and pleading for the research community to change course.^35^

On the other hand, in our optimistic scenario, we evaluated the implications of relying only on the EMERGE trial (and assuming equivalent efficacy in mild AD patients) to reflect current knowledge as suggested by the manufacturer. While the rationale for this approach has been convincingly critiqued^12-14,36^, we note that, perhaps counterintuitively, it actually weakens the case for conducting another trial (NoTrt/NoTrial is optimal for WTP≤$200K) and thus CED. This is because, at the current price, even at WTP=$200K, the cost-effectiveness threshold is HR≤0.51, a value outside the 95% confidence interval for the EMERGE estimate that was used to derive an uncertainty distribution for the HR. In contrast, the meta-analytic predictive distribution – meant to reflect our uncertainty about the true effect for the target population (US MCI and mild AD patients who might take aducanumab) - incorporates between-trial heterogeneity and thus exhibits greater dispersion around the point estimate. There is therefore a higher, albeit small, chance in the latter case that the true benefit from aducanumab is large enough to justify its current cost.

### Limitations

We discuss additional limitations of our modeling approach in the Appendix.

## Conclusions

Our results confirm those of ICER and justify CMS’s decision not to cover aducanumab as treatment for the general population of beneficiaries with early-stage clinical AD. Moreover, our results quantify the magnitude of opportunity loss ($10B-$110B) in monetized societal health-related welfare that would have been expected in the US if CMS had reversed course.

Conducting an RCT to further evaluate the clinical impact of high-dose aducanumab would be expected to reduce uncertainty for clinicians and policymakers. However, whether and to what extent such a trial (and thus CED) would be expected to have a positive net societal benefit in the US depends primarily on how CMS would incorporate the trial results into its coverage decision and, to a lesser extent, our societal WTP for a QALY. Assuming CMS policymakers will ultimately be able to avoid the pitfalls of a legal framework that limits their ability to explicitly consider the opportunity costs of healthcare-directed resources when making coverage decisions, we conclude that the CED decision (with ENVISION) is at least reasonable, if not optimal, if a QALY is valued ≥$150K and the EMERGE and ENGAGE trial results are both taken at face value and relied on exclusively to represent current knowledge. The case for future research would become less ambiguous, however, if the manufacturer again voluntarily dropped the price significantly.

## Data Availability

All data produced in the present study are available upon reasonable request to the authors

