## Appendix for "Was CED the Right Choice? A Decision-Theoretic Evaluation of CMS’s ‘Cover with Evidence Development’ Policy for Aducanumab"

**Supplementary Methods**

*Model*

The entire cohort is instantiated at an initial age and progressively ages each cycle (depending on cycle length). Separate instances of each severity state are used for different combinations of sex, setting – long-term care (LTC) vs. community dwelling – and treatment status – no treatment, treatment, and discontinued treatment. Transitions are allowed from community to LTC at all AD severity levels, but patients cannot return to the community setting. While the risk of transitioning to LTC increases with AD severity, transitions between disease severity states are assumed to be constant across settings. Moreover, conditional on surviving a cycle, transitions across AD severity states and to LTC are assumed to be age and sex invariant.

Probabilities governing transitions between alive states are parameterized as conditional on surviving the cycle. Point estimates for the annual (conditional) transition probabilities between AD severity states (in a natural history setting) are given as a transition matrix in eTable 2. To transform transition probabilities to a shorter cycle length, we sought to calculate an appropriate matrix root of the transition matrix.^1^ However, the desired exact root matrices were found not to be proper stochastic matrices which suggests the process characterized by this transition matrix is not embeddable in an underlying continuous-time Markov process.^2^ We therefore used the Shur-Padè method to select an optimal approximate root stochastic transition matrix, where optimality is defined with respect to a distance measure placed on residuals on the annual scale.^2,3^

We assume patients undergo treatment with high-dose Aducanumab (titrated to 10 mg/kg of bodyweight) while in the MCI or mild AD state (as per the FDA label). The effect of Aducanumab is implemented in the model via a HR on the baseline hazard rate of progressing to any more advanced AD state. The probabilities of transitioning to specific more advanced states, e.g., from MCI to mild vs. moderate AD, are then renormalized to account for the new (reduced) probability of progressing at all. We assume the probability of regressing to a less severe AD state is not impacted; rather, Aducanumab alters the probability of remaining in the current state. Point estimates for the annual (conditional) transition probabilities between AD severity states while on treatment are given as the first two rows of a transition matrix in eTable 3.

Patients are assumed to receive the full effect (altered transition rates) throughout any cycle they take the drug, including the first cycle (during titration) and a cycle in which they discontinue treatment due to a symptomatic adverse event. However, beginning the next cycle, Aducanumab is assumed to have no effect on patients who have stopped taking the drug due to symptomatic adverse events or progression to moderate AD. We justify these modeling decisions by noting that the HR estimate we use is derived from a time-invariant and intention-to-treat analysis and thus averages over time-varying effectiveness of the drug, e.g., delay in benefit due to titration, and patient discontinuation, respectively. In the absence of data, we assume the treatment effect is common across age, sex, and setting.

For our primary analyses, we derive an estimate of the treatment HR for MCI patients by performing a Bayesian meta-analysis^4^ of the EMERGE and ENGAGE trial results. While the relevant estimate is available for the EMERGE trial^5^, the manufacturer has not shared or published an estimate of the HR for the ENGAGE trial. We therefore relied on an estimate constructed by ICER researchers from available trial results.^6^ Similarly, in the absence of manufacturer-reported estimates of the treatment HR for mild AD patients, we again followed ICER in assuming Aducanumab is half as effective in this population (50% reduction on the log-HR scale). However, following ICER^6^ and a modeling study funded by the manufacturer^5^, we also considered an optimistic scenario using only the EMERGE trial HR for MCI patients and assuming equal efficacy among mild AD patients.

The target population for our analyses is the entire US population with beta-amyloid positive MCI due to AD or mild AD who would take Aducanumab or not, depending on the coverage decision. It is unlikely that the results from the actual trials would generalize perfectly to the average effect for this target population due to differences in disease severity, age, comorbidities, treatment implementation, and other unknown factors.^7^ Thus, we use our meta-analytic model to quantify uncertainty about what the true effect would be in the target population.^8^ In particular, we model parameter uncertainty for the MCI HR using an empirical distribution derived from thinned samples from the meta-analytic posterior predictive distribution for a new trial, allowing for between-trial heterogeneity. Given the difficulty of estimating the degree of between-trial heterogeneity in a meta-analysis with only two trials, we placed an (empirically informed) informative prior on the heterogeneity variance.^9^ To capture the extreme uncertainty associated with the mild AD HR, a (0,1) uniform distribution was placed on the relative efficacy parameter (proportion by which treatment efficacy is reduced).

Patients in all states face an elevated risk of death attributable to AD compared with the appropriate general US population age- and sex-specific mortality risk. Similarly, patients are assumed to suffer an AD-attributable decrement to health-related quality of life (HRQoL). The increased mortality risk is parameterized as a hazard ratio (HR) while the HRQoL decrement is assumed to be additive. Both the mortality HR and HRQoL decrement increase with AD severity but are assumed to be age-invariant. Ignoring adverse events, Aducanumab is assumed to only indirectly affect mortality and HRQoL by altering transition rates between severity states. For age- and sex-specific baseline mortality rates and age-specific baseline HRQoL weights, the model uses lifetables from the Human Mortality Database^10^ and general US population HRQoL weights (see eTable 1 for details and eTable 4 for values), respectively.

For analyses undertaken from the US healthcare perspective, only patient HRQoL and QALYs are considered, but, for the societal perspective, the sum of caregiver and patient QALYs is used to account for the substantial impact providing routine care to a patient with AD has on a caregiver’s daily life and quality of life.^11^ Like patient HRQoL, caregiver HRQoL decreases with age and is diminished due to patient AD status, depending on severity. We assume each patient has one caregiver of the same age. For simplicity, caregiver mortality is not considered, and the HRQoL decrement for caregivers paired with a deceased patient is set equal to that of severe AD. While perhaps counterintuitive, this approach avoids exaggerating the benefit of a (patient) life-prolonging intervention by appearing to extend the life of a caregiver.

Following ICER^6^ and in line with reported data^12^, we assume, within a year of treatment induction, 10% of patients will experience a symptomatic adverse event in the form of amyloid-related imaging abnormalities (ARIA) necessitating permanent discontinuation of treatment. In the model, these patients face a temporary HRQoL decrement but no increased mortality risk.

For the healthcare perspective, the model includes treatment-related and unrelated medical costs and LTC costs. Treatment-related medical costs include drug acquisition (accounting for waste) and administration (via IV in an outpatient setting) costs and MRI scans of the brain to detect and monitor ARIA. Unrelated medical costs are incorporated by applying AD-severity-specific (but age-invariant) cost multipliers to age-specific average annual medical costs for the general US population. The model also includes the costs of drugs (donepezil and memantine) used to treat symptoms in non-severe AD patients. LTC costs are based on residence in a skilled nursing facility.

For the societal perspective, the model also includes patient and caregiver time and transportation costs associated with receiving treatment and undergoing monitoring, patient productivity costs, and the cost of labor for informal caregiving. For these costs, each hour of time or labor is valued at the average hourly wage for private-sector compensation in the US. For productivity costs, in the reference case, we assume all patients (a population 70+) would produce 20 hours a week of formal or informal labor or household production in the absence of AD, with each hour valued as above. We further assume the proportion of patients unable to produce this labor increases with AD severity. We note that, while the model includes survivor medical costs – those that accrue in extended life years – and survivor productivity costs, we exclude survivor consumption costs because of consistency concerns.^13^ Full details about specific costs included, values used, uncertainty distributions (if any), and evidence sources are given in eTable 5.

Here we highlight several differences between ICER’s model and our model. First, in its reference case, ICER assumed patients with moderate AD would continue receiving Aducanumab, and thus racking up costs, although without benefit. As per the FDA label, our model assumes patients stop taking Aducanumab when they transition to moderate AD. Second, ICER subtracted a caregiver HRQoL decrement (attributable to AD) from patient HRQoL and thus only included modified patient QALYs in their analyses from the societal perspective. We believe ICER’s approach would undervalue treatment-related extension of life as the gain in QALYs in the model would be smaller than the patient’s actual gain. Our model sums patient and caregiver QALYs, maintaining a constant caregiver HRQoL once a patient dies (to avoid overcounting life extensions). Third, our model uses the recently updated drug acquisition price (50% reduction). Fourth, we use different (larger) severity-state-specific cost-multipliers for background unrelated medical costs taken from what we believe is a more nationally representative evidence source.^14^ Fifth, unlike ICER’s model, we do not include caregiver direct medical costs as we deemed the evidence supporting a link between providing care for AD patients and increased medical costs to be inconclusive.^15^ Sixth, we included patient and caregiver time and transportation costs associated with receiving treatment in the societal perspective while ICER did not.

Finally, we took a different approach to productivity costs. In ICER’s analyses from the societal perspective, they counted only lost hours of formal (market) labor attributable to AD among MCI and mild AD patients who would otherwise be working more. This has an intuitively problematic effect of decreasing productivity costs as a patient’s disease progresses to moderate and severe AD. ICER’s rationale was that more advanced AD patients in its data source did not work and it was implausible that moderate and severe AD patients would work. However, we believe this is an illusion created by selection bias since none of the patients are able to work and thus none report reduced hours in the data source used by ICER. However, in the absence of AD, they would be able to produce labor, be it formal, informal or household production. Moreover, although no formal recommendation was made, the 2^nd^ panel on Cost-effectiveness acknowledged the need to consider informal labor and household production among elderly/retired populations.^8^

The model underwent extensive internal validation and was validated against ICER’s results. In particular, for both treatment and control, we compared point estimates of time spent in each health state, LYs (both overall and spent in the community), and QALYs and lifetime costs from the healthcare sector perspective. We also compared incremental (treatment minus control) LYs, QALYs, and costs, and point estimates of the incremental cost-effective ratio, defined as the ratio of incremental costs (LYs or QALYs) to incremental effects. Finally, for the treatment arm, we compared the proportion of the cohort still receiving treatment and alive each year and lifetime Aducanumab acquisition costs. For these validation exercises, we used ICER’s modeling choices and parameters whenever possible.

We also performed several sensitivity analyses to assess the impact of potentially important modeling choices on our results. In particular, we informally evaluated the impact of different choices on our cost-utility results. For choices deemed potentially influential at the decision level or very influential with respect to magnitude, we performed a sensitivity analysis on our VOI results. We considered the following scenarios: (1) excluding caregiver QALYs from the societal perspective, (2) using ICER’s severity-state-specific cost multipliers on background medical costs, (3) excluding patient and caregiver time and transportation costs associated with treatment from the societal perspective, (4) valuing caregiver informal labor at the average hourly total compensation for a home health aide (as per recommendations^8^), and (5) using ICER’s approach to productivity costs.

*Modeled Cohort*

Following ICER, for the cost-utility and per-person VOI modeling analyses, we used a mixed cohort of MCI (55%) and mild AD (45%) patients primarily in a community setting (92%). The former breakdown (55% vs. 45%) is derived from an analysis^16^ of a population of beta-amyloid positive AD patients while the latter (92% vs. 8%) is an estimate of the distribution of living setting for 65-74 year old AD patients.^17^

*Per-person EVSI*

While the gold standard for estimating the per-person expected value of a decision after reducing uncertainty in a subset of parameters, i.e., the first term in Equations 2-3, involves nested Monte Carlo sampling with embedded Markov Chain Monte Carlo for Bayesian updating, this process is generally very computationally expensive, often prohibitively so.^18^ We therefore used established approximation methods based on generalized additive models to estimate the per-person EVSI^19^. Moreover, to limit the dimension of the considered parameter space and thereby maintain the feasibility and validity of this approximation method, we did not include adverse-event related parameters in our calculations even though a future trial would certainly provide such information. However, early exploratory analyses suggested this approach to be reasonable.

To calculate per-person EVSI, a study design, population, and simulation implementation must be specified. The study population and considered designs are discussed in the main text; here we briefly describe simulation implementation. Within the trial, patient transitions from MCI to mild AD and from mild to moderate AD are simulated from treatment-arm-specific exponential distributions based on the model’s homogenous annual transition probabilities. We assumed that MCI patients who progress to mild AD during the trial would in general remain on treatment and continue to be followed-up. Similar to the completed trials, we assume 10% of patients would withdrawal from follow-up within 18 months and 5 in 1,000 would die. Censoring and death times, among those experiencing the event, were simulated using a truncated gamma and continuous uniform distribution, respectively.

*Population EVSI*

In the calculation of population-level EVSI (Equation 5) the per-person value is discounted (if at all) over and above any discounting applied in the calculation of the per-person measure although at the same rate. We note that for population EVSI the value of information accrues only after a trial is completed, and so an assumption about the time to trial results is needed.^20^ This requires making an assumption about the delay from the present to randomization to treatment (including an enrollment period) and the delay from the end of follow-up to the public presentation, e.g., publication, of results. We assumed a 12-month delay to randomization, and a 12-month delay from the end of follow-up to results. The latter is the maximum allowable time for a CMS approved trial under its preliminary CED decision.^21^ The sum of these terms, along with the follow-up period, determines the time from the present to the availability of results, i.e., the time at which the information begins to have decision theoretic value.

*Trial Costs*

We note that the opportunity costs of a trial (Equation 6) could be negative, i.e., a cost-savings, if the initial coverage decision is not made based on the sign of the INMB. The cost of treating (over and above research costs) the other half of trial participants – those receiving the status quo treatment – is not included because such patients would receive identical treatment in the absence of a trial.

To garner face validity, an estimate of the expected direct research costs of a RCT should vary with the number of trial participants, the length of follow-up, and the intensity of follow-up required. To satisfy this criterion, we derive an estimate of total expected direct research costs from an estimate (based on 225 trials) of the cost per patient visit to a research clinic in a recent and typical RCT evaluating a novel therapeutic agent.^22^ We perform this calculation under three scenarios, using the estimated median ($3,934), lower quartile ($2,818), and upper quartile ($5,869) cost per-patient-visit (with values inflated to 2020 US dollars using the personal consumption expenditure, or PCE, price index). We assume that, similar to the initial phase III trials, patients enrolled in a future trial would undergo a visit at baseline, 4 visits in the first year, and then 2 visits every 6 months. Consequently, the expected direct research costs (Equation 7) of a future trial evaluating Aducanumab will vary with the sample size and length of follow-up.

*Power Calculation*

In order to compare our VOI results with the usual methods of trial design and to inform our analysis of the expected consequences of conducting a trial like the manufacturer’s planned ‘ENVISION’ trial, we performed a simulation-based power analysis. In particular we performed a set of sample size calculations using the same trial design, follow-up, population, and estimator (unadjusted Cox proportional hazard model) as with the EVSI. We assumed the trial would target 90% power (two-sided test with a 0.05 false-positive rate) to detect a benefit for MCI patients. We performed the calculation assuming a true MCI HR of 0.69 and 0.84 (and a true relative effectiveness of 1 and 0.5 for mild AD patients), corresponding to the EMERGE/optimistic and meta-analytic/ICER point estimates, respectively. For comparison, we repeated the analyses using analytic methods based on proportional hazards.^23^

*Population INMB*

Equations 8-10 (below) define the population-level expected INMB of the three available coverage and trial strategies (in addition to don’t cover, no trial) among patients who might initiate aducanumab in the next 15 years. NMB is defined as monetized health-related welfare net costs, with health-related welfare measured in terms of quality-adjusted life years (QALYs). QALYs are converted to dollars based on the societal willingness to pay (WTP) for one QALY. Costs include those associated with treatment as well as any trial. For technical reasons related to our analytic methodology, we report INMB, i.e., net that of a reference alternative (deny coverage, no trial). Thus, by definition, the INMB of option 1 (deny coverage, no trial) is 0.

eTable 1: Model Parameters, Uncertainty, and Evidence Sources

| **Parameter** | **Value** | **Uncertainty Distribution** | **Data Source** |
| --- | --- | --- | --- |
| AD Natural History Parameters | | | |
| Annual transition probabilities between disease stages conditional on surviving the year in a natural history setting. | See eTable 2 | Multivariate truncated normal distribution for each state with dimension k-1, where k is the number of probabilities > 0. Covariance parameters derived assuming data generated from multinomial distribution. | NACC (UDS)^16^ |
| Annual disease-stage-specific probabilities of transitioning from a community setting to LTC | MCI = 0.024  Mild AD = 0.038  Moderate AD = 0.11  Severe AD = 0.259 | Marginal beta-distributions correlated via a Gaussian Copula distribution with exchangeable correlation | CERAD^24^ for AD; assumption made by ICER for MCI^6^ |
| AD-attributable HR on general US population age- and sex- specific annual average all-cause mortality hazard rates | MCI = 1.82  Mild AD = 2.92  Moderate AD = 3.85  Severe AD = 9.52 | Multivariate normal distribution on log-HR based on 95% CIs and assumed correlation | Population-based study in Denmark^25^ |
| General US population age-and sex- specific annual average all-cause mortality hazard rates | Sex-specific life tables calculated for each age over a 5-year period (1x5 table: 2014-2019) | None | The Human Mortality Database^10^ |
| Treatment Efficacy Parameters | | | |
| Aducanumab-attributable HR on the rate at which patients in the MCI state progress to more advanced disease | HR=0.84 | An empirical distribution derived from thinned (independent) samples from the meta-analytic posterior predictive distribution for the log-HR of a new trial.  Mean=-0.174;  Stdev=0.298 | Bayesian re-analysis of a Meta-analysis^5,6^ of EMERGE & ENGAGE Trials |
| Relative size of the benefit of Aducanumab on Mild AD patients compared with patients in the MCI state | 50%  (implies HR=0.92) | Uniform distribution over  (0, 1) | Assumption taken from ICER^6^ based on EMERGE & ENGAGE Trials |
| Treatment AE Parameters | | | |
| Probability of any type of treatment-induced ARIA | 0.31 | Normal distribution based on 95% CI (0.28, 0.35) | Manufacturer-funded modeling analysis^5^ |
| Probability of treatment-induced symptomatic ARIA resulting in a HRQoL decrement | 0.10 | Normal distribution based on 95% CI (0.08, 0.12) | FDA briefing document^12^ |
| Mean duration of ARIA event (asymptomatic and symptomatic) | 12 weeks | Continuous uniform distribution over 9-18 weeks | FDA briefing document^12^, uncertainty distribution based on assumption |
| Probability of discontinuing treatment due to AE (within 1 year) | 0.10 | Continuous uniform distribution over (0.08-0.12) | FDA briefing document^12^, uncertainty distribution taken from ICER |
| HRQoL Parameters | | | |
| General US population age-specific HRQoL weights | Cubic spline interpolation and extrapolation of median preference weights for 10-year age categories. See eTable 4 | Multivariate normal distribution based on reported standard errors assuming exchangeable (0.75) correlation. | Preference weights^26^ mapped from EQ-5D scores from the nationally-representative MEPS survey (2000-2002)^27^ |
| AD-attributable HRQoL decrements for the patient (severity- and setting- specific) | Community  MCI = -0.17  Mild AD = -0.22  Moderate AD = -0.36  Severe AD = -0.53  LTC  MCI = -0.17  Mild AD = -0.19  Moderate AD = -0.42  Severe AD = -0.59 | Multivariate truncated normal distribution (truncated at +/- 3 standard deviations) with exchangeable (0.75) correlation. Standard deviations taken from ICER report^6^. | Taken from ICER’s report^6^; based on a cross-sectional study^11,28^ of AD patients; used HUI:2 and (standard gamble derived) community preference weights. |
| AD-attributable HRQoL decrements for the caregiver (severity- and setting- specific) | Community  MCI = -0.03  Mild AD = -0.05  Moderate AD = -0.08  Severe AD=Dead=-0.10  LTC  MCI = -0.03  Mild AD = -0.05  Moderate AD = -0.08  Severe AD=Dead=-0.10 | Multivariate truncated normal distribution (truncated at +/- 2 standard deviations) with exchangeable (0.75) correlation. Standard deviations set at 50% of the mean. | Same as above. |
| ARIA utility decrement  (for duration of symptomatic ARIA) | -0.14 | Uniform distribution  (-0.20, -0.06) | ICER report^6^  (HRQoL weight for a headache^29^) |
| Expected annual utility decrement from other AE (among all treated patients) | -0.0003 | Uniform distribution  (-0.0006, 0) | Manufacturer-funded modeling analysis^5^ |

Abbreviations: AD – Alzheimer’s Disease, NACC – National Alzheimer’s Coordinating Center, UDS – Unified Data Set, LTC – Long-Term Care, MCI – Mild Cognitive Impairment, CERAD – Consortium to Establish a Registry for Alzheimer’s Disease, HR – Hazard Ratio, CI – Confidence Interval, Stdev – Standard Deviation, ICER – Institute for Clinical and Economic Review, AE – Adverse Events, ARIA – Amyloid Related Imaging Abnormalities, HRQoL – Health Related Quality of Life, EQ-5D – EuroQol-5D, MEPS – Medical Expenditure Panel Survey, HUI:2 – Health Utilities Index Mark 2

eTable 2: Natural History Annual Transition Probabilities*

|  | MCI | Mild AD | Moderate AD | Severe AD |
| --- | --- | --- | --- | --- |
| MCI | 0.770 | 0.168 | 0.060 | 0.002 |
| mild AD | 0.033 | 0.571 | 0.349 | 0.047 |
| Moderate AD | 0 | 0.026 | 0.558 | 0.416 |
| Severe AD | 0 | 0 | 0.024 | 0.976 |

Abbreviations: AD – Alzheimer’s Disease, MCI – Mild Cognitive Impairment

* Probabilities are conditional on surviving the year and represent point estimates (most likely values)

eTable 3: Annual Transition Probabilities with Aducanumab*

|  | MCI | Mild AD | Moderate AD | Severe AD |
| --- | --- | --- | --- | --- |
| MCI | 0.803 | 0.144 | 0.051 | 0.002 |
| mild AD | 0.033 | 0.597 | 0.326 | 0.044 |

Abbreviations: AD – Alzheimer’s Disease, MCI – Mild Cognitive Impairment

* Probabilities are conditional on surviving the year and represent point estimates (most likely values) for both natural history transition probabilities and the (meta-analytic) effect of Aducanumab. It is assumed that Aducanumab only affects transitions from MCI and mild AD.

eTable 4: Baseline US Adult Population HRQoL Weights by 10-year Age Categories*

| Age-Range | Assumed (Mean) Age for Interpolation† | Mean HRQoL (Stderr) | Median HRQoL¶ |
| --- | --- | --- | --- |
| 18-29 | 24 | 0.922 (0.0019) | 1.00 |
| 30-39 | 35 | 0.901 (0.0021) | 1.00 |
| 40-49 | 44 | 0.871 (0.0024) | 0.844 |
| 50-59 | 54 | 0.842 (0.0028) | 0.827 |
| 60-69# | 64 | 0.823 (0.0034) | 0.827 |
| 70-79# | 74 | 0.790 (0.0036) | 0.810 |
| 80+# | 84 | 0.736 (0.0062) | 0.778 |

Abbreviations: HRQoL – Health-Related Quality of Life, Stderr – Standard Error

Used earlier ages to fit cubic interpolating spline. But only 70+ was relevant

* Values are derived from mapping EQ-5D scores from the nationally-representative MEPS (2000-2002) survey^27^ to preference weights (0-1) estimated from a US community sample^26^

#Implies age-rage was directly relevant for the model. Earlier data points were used to fit an interpolating cubic spline that was then used to extrapolate to 100.

†Uncertainty of the mean age in each category was incorporated into the PSA

¶ The model uses a HRQoL curve fit to Median values.

eTable 5: Costs: Values, Uncertainty, and Evidence Sources.

| **Parameter** | **Value*** | **Uncertainty Distribution** | **Data Source** |
| --- | --- | --- | --- |
| Drug Acquisition and Administration Costs | | | |
| First year mean acquisition cost (assuming no waste) | $20,500  (previously $39,000) | None | Recent manufacturer estimate based on a 50% reduction in acquisition price^30^ |
| Mean annual acquisition cost (assuming no waste) after year 1 | $28,200  (previously $56,000) | None |  |
| Assumed vial waste | 6%  (above prices multiplied by 1.06) | None | ICER^6^ Assumption |
| Cost per IV administration session | $74.58 | None | Medicare reimbursement rate^31^ (HCPCS Code 96365) |
| Frequency of drug administration | Every 4 weeks | None | Treatment protocol from trials |
| Costs Related to Monitoring and Adverse Events | | | |
| Cost per brain MRI Scan to detect new ARIA and/or monitor ongoing ARIA | $255.33 | None | Medicare^31^ reimbursement rate (HCPCS Code 70553) |
| Number of brain MRI scans in the first year and in later years | 1^st^ year = 3  2+ years = 0 | None | Treatment protocol from trials |
| Frequency of brain MRI scans for patients with ARIA | Every 4 weeks | None | FDA briefing document^12^ |
| Non-Treatment-Related Medical Costs | | | |
| US population age-specific mean annual medical costs | See Appendix table | Normal marginal distributions for each age based upon 95% CI’s. Assume perfect correlation across ages to preserve ordering. | An analysis of the nationally representative MEPS survey (2007-2015)^32^ |
| AD severity-specific medical cost multipliers (vs. cognitively normal adults) | MCI = 1.63  Mild AD = 2.32  Moderate AD = 2.54  Severe AD = 2.54  [REFERENCE CASE] | Bivariate normal distribution on the log cost ratios (MCI and Mild AD) with exchangeable correlation of 0.75 | 2 Years after diagnosis of MCI or mild AD in 5% sample of Medicare beneficiaries (2009-13)^14^; assumption for mod/severe |
|  | MCI = 1.13  Mild AD = 1.59  Moderate AD = 1.69  Severe AD = 1.69  [Sensitivity Analysis] | Multivariate normal distribution on the log cost ratios with exchangeable correlation of 0.75 and ad hoc exclusion of parameter sets violating natural ordering | Comparison of 1 year of costs for random population sample of Olmstead County 70+ residents.^33^ Adjusted for age, sex, an education |
| Daily cost of donepezil (10 mg) and memantine (10 mg) given to 1/3 of mild AD and 1/3 of moderate AD patients once and twice daily, respectively | Donepezil = $0.22  Memantine = $0.68 | None | As included in the ICER^6^ model based on an analysis of Medicare claims (2008-16)^34^ |
| Long-Term Care Costs | | | |
| National Average monthly costs attributable to residence and care in a skilled nursing facility | $7,186 | None | Administration on Aging^35^ (2016), adjusted to 2020 by ICER report^6^ |
| Treatment Time & Transportation Costs | | | |
| Transportation cost per visit for infusion or MRI | $15 | Continuous uniform distribution over $5-25 | Assumption |
| Time per visit for infusion or MRI | 2.5 hours (for patient and caregiver) | Continuous uniform distribution over 1.5-3.5 hours | Manufacturer report^12^ (hour per infusion) and assumption |
| Average hourly wage used to value an hour of patient and caregiver time | $29.47 | None | BLS^36^ |
| Productivity Costs | | | |
| Average hourly wage used to value an hour of lost patient productivity | $29.47 | None | BLS^36^ |
| Counterfactual (no AD) number of hours of formal or informal labor or household production per week per patient  [Reference case] | 20 | Continuous uniform distribution over 10 to 30. | Assumption (between 0 and 40) |
| Proportion of patients in the formal labor market  [Sensitivity Analysis] | MCI = 0.214  mild AD=0.094  moderate AD = 0  Severe AD = 0 | bivariate truncated normal distribution with an assumed 0.5 exchangeable correlation and marginal moments based on binomial proportion asymptotic theory | GERAS-US Study^15^  (prospective cohort) |
| Proportion patients unable to undertake (any) formal or informal labor or household production because of AD | MCI = 0.032;  mild AD=0.138  moderate AD=0.75  severe AD = 1 | bivariate truncated normal distributions with an assumed 0.5 exchangeable correlation and marginal moments based on binomial proportion asymptotic theory;  For moderate AD, continuous uniform distribution 0.5-1 | GERAS-US Study^15^  (prospective cohort) and assumption for moderate and severe AD. |
| Number of hours of formal labor lost/week by patients among those reducing work due to disease.  [Sensitivity Analysis] | 20 | Continuous uniform distribution over 10 to 30. | Assumption made in ICER report^6^ |
| Informal Caregiver Costs | | | |
| Average hourly wage used to value an hour of informal caregiving | $29.47  [Reference Case] | None | Average hourly private-sector compensation:  BLS^36^ |
|  | $17.67 (mean wage=$13.49; assumed fringe rate=0.31)  [Sensitivity Analysis] | None | Average hourly compensation for a home health aide: BLS^37^ |
| Mean number of hours of informal caregiving required per month for AD patients living at home (by disease severity) | MCI = 69.2  Mild AD = 113.0  Moderate AD = 169.1  Severe AD = 297.8 | Multivariate normal distribution with assumed 0.5 exchangeable correlation. Marginal standard deviations ranged 7-10. | MCI from GERAS-US study^15^ and AD’s from GERAS-Europe^38^ |
| Proportionate reduction of informal caregiving hours needed per month for AD patients living in a skilled nursing facility | 0.44 | Continuous uniform distribution over 0.29 to 0.6 | Assumption made in ICER report^6^ |

Abbreviations: ICER – Institute for Clinical and Economic Review, WAC – Wholesale Acquisition Cost, HCPCS – Healthcare Common Procedure Coding System, ARIA – Amyloid Related Imaging Abnormalities, CI – Confidence Interval, MEPS – Medical Expenditure Panel Survey, AD – Alzheimer’s Disease, MCI – Mild Cognitive Impairment, BLS – Bureau of Labor Statistics

*All dollars values are given in 2020 US Dollars.

*Equations*

Equation 1 defines the per-person expected incremental net monetary benefit (INMB_per_), with expectation explicitly taken over uncertainty in the model parameter vector, $\theta$, and implicitly over individuals. The subscripts Trt and NoTrt refer to Aducanumab and treatment as usual, respectively. The term $\lambda$ is the monetary value placed on a single QALY.

$E_{\theta}\left[ {INMB}_{per}|\theta\right]=E_{\theta}\left[ \left( \left( QAL{Ys}_{Trt}-QAL{Ys}_{NoTrt} \right)\times\lambda-\left( Costs_{Trt}-{Costs}_{NoTrt} \right) \right)|\theta\right]$	(1)

Equation 2 defines the standard per-person expected value of sample information (EVSI_per_). It is defined as the difference between the expected value of a decision made after collecting sample information (with a given design) and the expected value of a decision made given current information. The terms $E_{\theta}\left( {INMB}_{per}|\theta\right)$ and $E_{\theta|X}\left( {INMB}_{per}|\theta\right)$ refer to the per-person expected INMB, averaging over uncertainty in the model parameter vector, $\theta$, given current information and after observing the results, X, of the trial, $\theta|X$, respectively. Collecting sample information is assumed to reduce uncertainty about some subset, $\psi$, of the full model parameter vector, $\theta$. In our case, we assume the relevant subset, $\psi$, represents the two treatment HRs (for MCI and mild AD).

${EVSI}_{per}=E_{X}\left( max\left\{ E_{\theta|X}\left( {INMB}_{per}|\theta\right), 0 \right\} \right)-max\left\{ E_{\theta}\left( {INMB}_{per}|\theta\right), 0 \right\}$	(2)

Equation 3 defines the per-person EVSI under the assumption that the decision made at present with current information, $D_{0}^{*},$ is based on an alternative unspecified decision criterion (rather than the sign of the per-person INMB) but the decision made after trial results are available, $D_{1}$, is based on the sign of the per-person INMB. Thus, only the second term of Equation 3 differs from Equation 2. The term $I_{\left\{ D_{0}^{*}=Trt \right\}}$ is an indicator variable set to 1 if the initial decision is to cover aducanumab and 0 for the opposite.

${EVSI}_{per}|D_{0}^{*}=E_{X}\left( max\left\{ E_{\theta|X}\left( {INMB}_{per}|\theta\right), 0 \right\} \right)-\left[ {I_{\left\{ D_{0}^{*}=Trt \right\}}*E}_{\theta}\left( {INMB}_{per}|\theta\right) \right]$	(3)

Equation 4 defines the per-person EVSI assuming both the decision made now with current information, $D_{0}^{*}$, and the decision made after collecting further information, $D_{1}^{'}$, are based on alternative decision criteria, with the latter assumed to depend on X. The term $I_{\left\{ D_{1}^{'}(X)=Trt \right\}}$ is an indicator variable analogous to that defined above.

${EVSI}_{per}|D_{0}^{*},D_{1}^{'}(X)=E_{X}\left[ I_{\left\{ D_{1}^{'}(X)=Trt \right\}}*E_{\theta|X}\left( {INMB}_{per}|\theta\right) \right]-\left[ {I_{\left\{ D_{0}^{*}=Trt \right\}}*E}_{\theta}\left( {INMB}_{per}|\theta\right) \right]$	(4)

Equation 5 defines the population-level EVSI (EVSI_pop_) in terms of the per-person EVSI (EVSI_per_). The term $\delta$ is the lag in months from the present to randomization to treatment, $\tau$ is the trial follow-up length in months, $\varepsilon$ is the delay in months from the end of follow up to the publication of results, and their sum is the time from present to trial results (when value starts accruing). The term floor() is the floor function. The term ${TrtN}_{t}$ is the number of patients in year t whose treatment would be affected by a revised decision (relative to current information) for years 1 to T, $T$ is the decision timeframe in years, and r is the discount rate.

${EVSI}_{pop}={EVSI}_{per}*\left( \sum_{t=(floor\left( \frac{\delta+\tau+\varepsilon}{12} \right)+1)}^{T} \frac{{TrtN}_{t}}{\left( 1+r \right)^{t-1}} \right)$	(5)

Equation 6 defines the expected total opportunity costs, $E[\left( {TC}_{Opp} \right)]$, of a future balanced RCT as a function of $D_{0}^{*}$ (the initial coverage decision), N (the total trial sample size, with N/2 per arm), and $\delta$ (the lag in months from the present to randomization to treatment). The term $I_{\left\{ D_{0}^{*}=NoTrt \right\}}$ is an indicator variable equal to 1 if the initial decision is to not cover Aducanumab and 0 otherwise (i.e., if the initial decision is to cover Aducanumab). The terms r and floor() are as above.

$E\left[ {TC}_{Opp}\left( D_{0}^{*} , N, \delta\right) \right]=\frac{\left( -1 \right)^{I_{\left\{ D_{0}^{*}=NoTrt \right\}}}*E_{\theta}\left( {INMB}_{per}|\theta\right)*\frac{N}{2}}{\left( 1+r \right)^{floor\left( \frac{\delta}{12} \right)}}$	(6)

Equation 7 defines the expected total direct research costs, ${E[TC}_{Res}]$, as a function of the cost per patient visit to a research clinic, $C_{PPV}$, N (total trial sample size, with N/2 per arm), $\delta$, the lag in months from the present to randomization to treatment, and $\tau$ the length of the follow-up period of the trial in months. The term ceil() denotes the ceiling function. $V_{t}$ is the number of visits per patient in year t (0 for $t\leq ceiling(\frac{\delta}{12})$). The $V_{t}$ terms depend on both $\tau$ and $\delta$. The term r is as above.

$E\left[ {TC}_{Res}(C_{PPV}, N,\tau, \delta) \right]=\left( \sum_{t=1}^{ceil\left( \frac{\tau+\delta}{12} \right)} \frac{V_{t}}{\left( 1+r \right)^{t-1}} \right)*N*C_{PPV}$	(7)

If we use $D_{trial}$ to denote the decision whether to conduct a trial (T$rial)$ or not $(NoTrial$), then the population-level expected INMB for the initial decision ($D_{0}^{*}$) to deny coverage without doing a trial, $E_{\theta}\left( {INMB}_{pop}|\theta, D_{0}^{*}=NoTrt, D_{trial}=NoTrial \right)$, is equal to 0. Equation 8 defines the

population-level expected INMB (over a T-year timeframe) for the strategy of offering coverage without a trial. The term ${TrtN}_{t}$ is the number of patients in year t whose treatment decision would be affected by the initial coverage decision for years 1 to T, and r is the discount rate

$E_{\theta}\left( {INMB}_{pop}|\theta, D_{0}^{*}=Trt, D_{trial}=NoTrial \right)= E_{\theta}({INMB}_{per}|\theta)*\left( \sum_{t=1}^{T} \frac{{TrtN}_{t}}{\left( 1+r \right)^{t-1}} \right)$	(8)

Equation 9 defines the population-level expected INMB (over a T year timeframe) for the strategy of denying coverage initially but conducting the optimal trial (assuming a trial is optimal) with sample size $N^{'}$ and follow-up length $\tau^{'}$ in months. For WTP values for which no trial is optimal, equation 9 reduces to 0. The equation can be broken down into three components (each line): the expected population-level INMB prior to (line 1) and after (line 2) the release of trial results and trial costs (line 3), which are subtracted from the sum of the first two components. The expected per-person INMB without treatment is 0. The sum in line 1 gives the total number of patients in the population effected by the initial treatment decision. The term $\delta$ is the lag in months from the present to randomization to treatment, $\tau^{'}$ is the optimal trial follow-up length in months, $\varepsilon$ is the delay in publishing results once follow-up is completed. Their sum is the time from present to the release of trial results. The term ${TrtN}_{m}$ is the number of patients in month *m* whose treatment decision is affected by the initial coverage decision. The expectation (first term) in line 2 is the *a priori* per-person expected INMB for a decision made (based on the sign of the INMB) after conducting the optimal trial. The sum (second term) in line 2 gives the total number of patients in the population affected by the post-trial decision. The third line is the expected total costs associated with the trial including the research costs (first term) and opportunity costs (second term) as defined in equations 7 and 6, respectively.

$E_{\theta}\left( {INMB}_{pop}|\theta, D_{0}^{*}=NoTrt, D_{trial}=Trial \right)=$	(9)

$$0 *\sum_{m=1}^{\delta+\tau^{'}+\varepsilon} \left( \frac{{TrtN}_{m}}{\left( 1+r \right)^{floor\left( \frac{\left( m-1 \right)}{12} \right)}} \right) +$$

$$E_{X}\left( max\left\{ E_{\theta|X}\left( {INMB}_{per}|\theta\right), 0 \right\} \right) *\sum_{m=\delta+\tau^{'}+\varepsilon+1}^{T*12} \left( \frac{{TrtN}_{m}}{\left( 1+r \right)^{floor\left( \frac{\left( m-1 \right)}{12} \right)}} \right) -$$

$${E[TC}_{Res}|N^{'},\tau^{'}]+E[\left( {TC}_{Opp}|{D_{0}^{*}=NoTrt, N}^{'} \right)]$$

Equation 10 defines the population-level expected INMB (over a T-year timeframe) for the strategy of offering coverage initially and conducting the optimal trial (assuming a trial is optimal) with sample size $N^{'}$ and follow-up length $\tau^{'}$ in months. For WTP values for which no trial is optimal, equation 10 reduces to equation 8. Equation 10 differs from equation 9 in two regards. First, the first term of line 1: the per-person expected INMB given current information associated with adopting treatment. Second, the expected opportunity costs associated with the trial (line 3) differ based on the initial coverage decision.

$E_{\theta}\left( {INMB}_{pop}|\theta, D_{0}^{*}=Trt, D_{trial}=Trial \right)=$	(10)

$$E_{\theta}({INMB}_{per}|\theta) *\sum_{m=1}^{\delta+\tau^{'}+\varepsilon} \left( \frac{{TrtN}_{m}}{\left( 1+r \right)^{floor\left( \frac{\left( m-1 \right)}{12} \right)}} \right) +$$

$$E_{X}\left( max\left\{ E_{\theta|X}\left( {INMB}_{per}|\theta\right), 0 \right\} \right) *\sum_{m=\delta+\tau^{'}+\varepsilon+1}^{T*12} \left( \frac{{TrtN}_{m}}{\left( 1+r \right)^{floor\left( \frac{\left( m-1 \right)}{12} \right)}} \right) -$$

$${E[TC}_{Res}|N^{'},\tau^{'}]+E[\left( {TC}_{Opp}|{D_{0}^{*}=Trt, N}^{'} \right)]$$

In both equations 9 and 10, for $m\in\left( \delta+1 \right):\left( \delta+6 \right)$, i.e., for the 6 months after the trial begins randomizing patients, ${TrtN}_{m}$ is adjusted to remove the $N^{'}$/2 patients randomized to the non-standard treatment, with $\frac{\left( N^{'}/2 \right)}{6}$ removed per month. These patients are accounted for in the trial opportunity cost quantity.

Equations 9 and 10 assume the post-trial coverage decision is made based on efficiency grounds, that is, the sign of the per-person expected INMB, as in the partially-generalized EVSI (equation 3). If, instead, the post-trial coverage decision is based on alternative decision criteria, $D_{1}^{'}\left( X \right),$e.g., the direction and statistical significance of the estimated effect, the first term in line 2 would be replaced by: $E_{X}\left[ I_{\left\{ D_{1}^{'}(X)=Trt \right\}}*E_{\theta|X}\left( {INMB}_{per}|\theta\right) \right]$, as in the fully-generalized EVSI (equation 4).

**Supplementary Results**

*Per-Person Cost-Utility Analysis*

eTable 6: Reference Case Modeled Outcomes for Societal and Healthcare Sector Perspectives

| Outcome | Untreated Cohort Mean  (95% CredInt) | | Treated Cohort Mean  (95% CredInt) | | Incremental  (Treated-Untreated)  Mean (95% CredInt)  & Probability>0 | |
| --- | --- | --- | --- | --- | --- | --- |
|  | r=0% | r=3% | r=0% | r=3% | r=0% | r=3% |
| Life Years | 6.737  (6.022, 7.555) | 5.883  (5.326, 6.515) | 6.917  (5.968, 7.985) | 6.014  (5.286, 6.816) | 0.180  (-0.498, 0.874)  Pr>0=0.78 | 0.131  (-0.383, 0.638)  Pr>0=0.78 |
| Quality-Adjusted Life Years (HCS) | 3.181  (2.703, 3.699) | 2.824  (2.427, 3.251) | 3.331  (2.605, 4.138) | 2.940  (2.345, 3.575) | 0.150  (-0.442, 0.749)  Pr>0=0.78 | 0.115  (-0.364, 0.586)  Pr>0=0.77 |
| Quality-Adjusted Life Years (Societal) | 23.258  (21.158, 25.377) # | 16.149  (14.956, 17.403) # | 23.434  (21.234, 25.707) # | 16.285  (14.969, 17.715) # | 0.176  (-0.512, 0.887)  Pr>0=0.78 | 0.136  (-0.417, 0.684)  Pr>0=0.77 |
| Lifetime Total Costs (HCS) in 2020 US Dollars | $346,000  ($276,000, $436,000) | $292,000  ($237,000, $362,000) | $457,000  ($366,000, $570,000) | $391,000  ($319,000, $479,000) | $112,000  ($60,000, $169,000)  Pr>0=1 | $98,000  ($58,000, $140,000)  Pr>0=1 |
| Lifetime Total Costs (Societal) in 2020 US Dollars | $743,000  ($612,000, $898,000) | $633,000  ($530,000, $752,000) | $857,000  ($720,000, $1,015,000) | $733,000  ($627,000, $853,000) | $114,000  ($77,000, $159,000)  Pr>0=1 | $100,000  ($76,000, $128,000)  Pr>0=1 |
| Incremental Cost-Effectiveness Ratio ($/QALY) Based on Posterior Means (HCS) † | --- | | | | $747,000 | $852,000 |
| Incremental Cost-Effectiveness Ratio ($/QALY) Based on Posterior Means (Societal) † | --- | | | | $648,000 | $735,000 |

Abbreviations: CredInt – 95% Posterior Credible Interval for the Mean, r – Discount rate, HCS – Health Care Sector Perspective, Societal – Societal Perspective, Pr - Probability

\# The arm-specific expected QALYs are not meaningful. Only the incremental values should be assessed. Because we do not model caregiver mortality, we assign a patient-disease related HRQoL-decrement to caregivers with deceased patients equal to that of severe AD. Thus caregivers continue to add age-adjusted HRQoL after their patients die. We do this to avoid exaggerating the benefit of extending a patient’s life which would occur if we were to set caregiver HRQoL to 0 when the patient dies.

† These values are calculated by dividing the posterior mean of the incremental costs by the posterior mean of the incremental QALYs.

eFigure 1: Per-Person Average INMB vs. WTP: Reference Case Analysis.

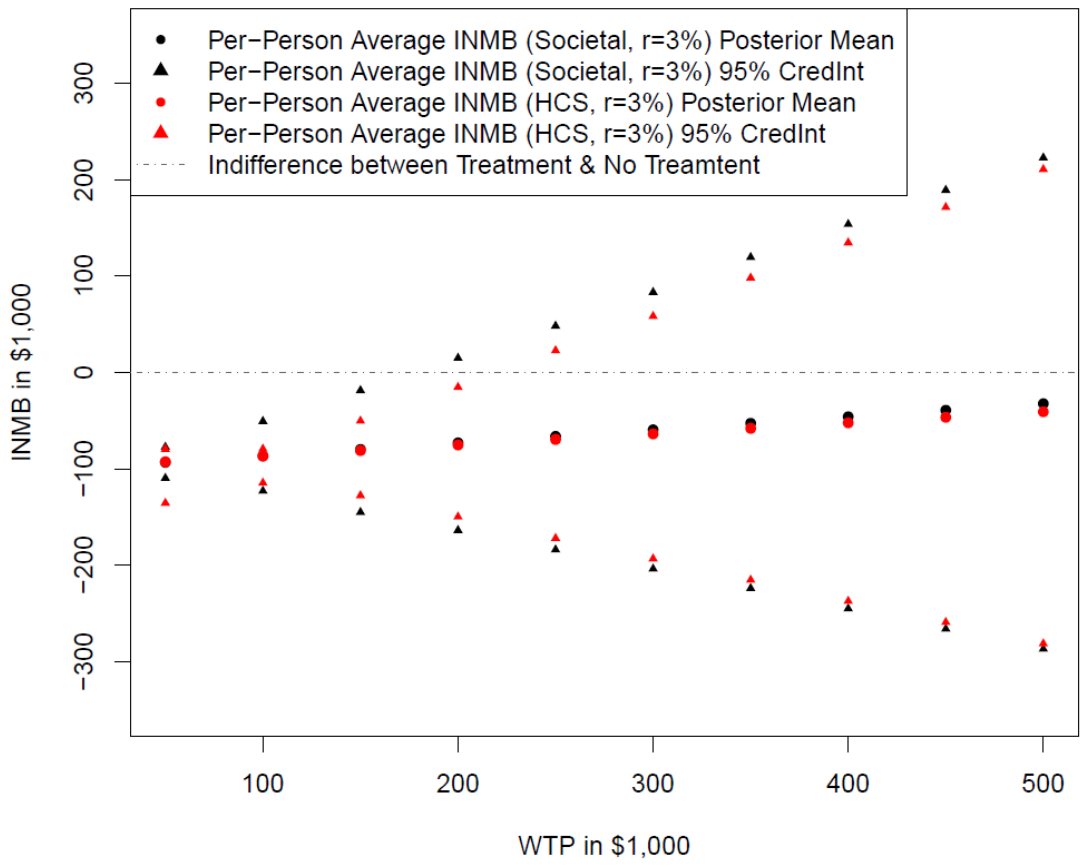

Abbreviations: INMB – Incremental Net Monetary Benefit, WTP – Willingness to Pay (for 1 quality-adjusted life year), Societal – Societal Perspective, HCS – Healthcare System Perspective, r – Discount Rate, CredInt – Credible Interval.

eFigure 1 displays the posterior mean and a 95% credible interval for the per-person average INMB from the societal and healthcare sector perspectives and when both costs and benefits are discounted at 3%. Treatment with aducanumab is optimal at a particular WTP given current information if and only if the posterior mean is greater than 0. The 95% credible interval limits depict our uncertainty about the true per-person average (across the population) INMB attributable to model parameter uncertainty.

eFigure 2: Cost-Effectiveness Acceptability Curve and Frontier for Reference Case

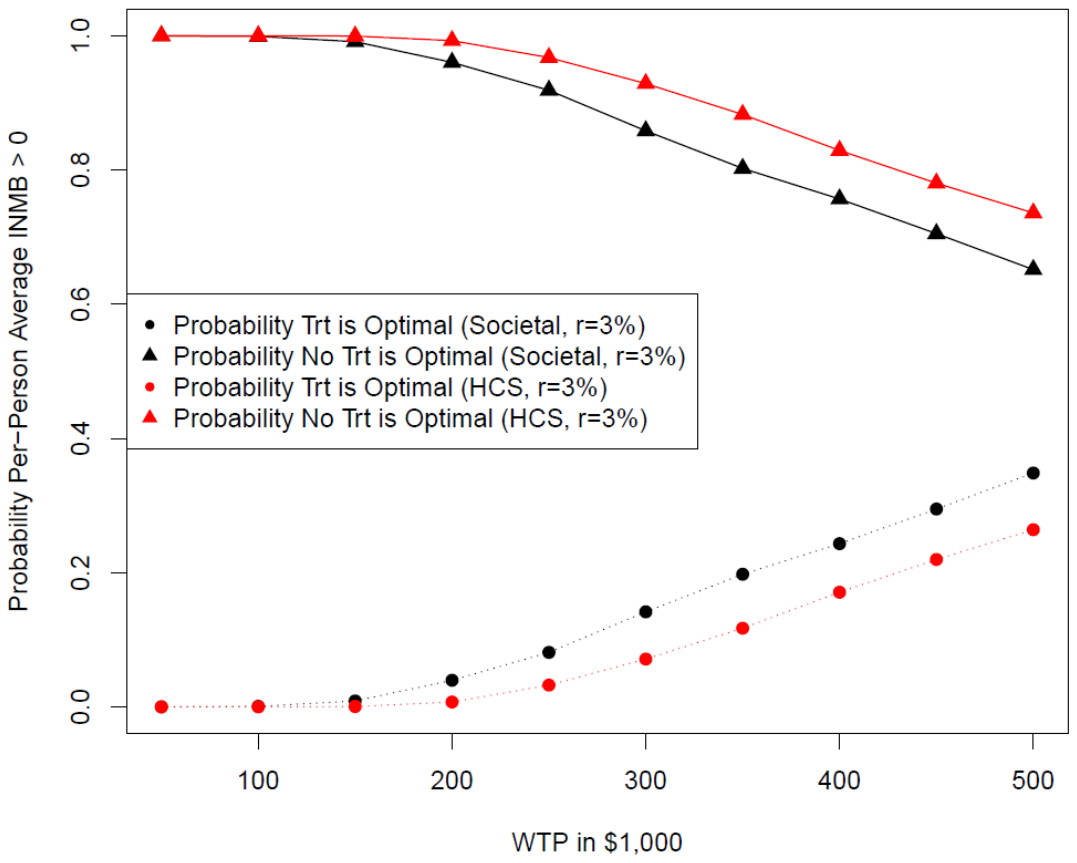

Abbreviations: INMB – Incremental Net Monetary Benefit, WTP – Willingness to Pay (for 1 quality-adjusted life year), Trt – Treatment (with Aducanumab), Societal – Societal Perspective, HCS – Healthcare System Perspective, r – Discount Rate.

eFigure 2 shows the probability treatment with Aducanumab (or no treatment) is optimal given current knowledge as a function of WTP for both the societal and healthcare sector perspective and when both costs and quality-adjusted life years are discounted at 3%. From an efficiency standpoint, treatment with Aducanumab would be optimal if and only if the per-person average INMB were greater than 0. However, because there is parameter uncertainty, this is not known with certainty (as is clear in eFigure 1). Thus treatment is optimal (given current knowledge) if and only if the expected per-person average INMB (displayed in eFigure 1 as the posterior means) is greater than 0, where the expectation is with respect to model parameter uncertainty. A solid line indicates the optimal decision using this criterion at each WTP while the dashed line indicates the suboptimal option.

eTable 7: Cost-Utility Analysis Results: Impact of Scenario Analyses

| Considered Scenario | Analysis  (r=3%) | Per-Person Average INMB: Posterior Mean# and (Probability > 0) | | | | |
| --- | --- | --- | --- | --- | --- | --- |
|  |  | WTP=$50K | WTP=$100K | WTP=$150K | WTP=$200K | WTP=$250K |
| Reference Case | Societal | -$93,000  (0) | -$87,000  (0) | -$80,000  (0.01) | -$73,000  (0.04) | -$66,000  (0.08) |
|  | HCS | -$93,000  (0) | -$87,000  (0) | -$81,000  (0) | -$75,000  (0.01) | -$70,000  (0.03) |
| Optimistic efficacy (EMERGE trial only) | Societal | -$116,000  (0) | -$94,000  (0) | -$71,000  (0) | -$49,000  (0.04) | -$27,000  (0.22) |
|  | HCS | -$125,000  (0) | -$106,000  (0) | -$87,000  (0) | -$68,000  (0) | -$48,000  (0.03) |
| Drug price reduced 50% ($14,100) | Societal | -$47,000  (0) | -$40,000  (0.04) | -$33,000  (0.13) | $-27,000  (0.23) | -$20,000  (0.32) |
|  | HCS | -$46,000  (0) | $-41,000  (0) | -$35,000  (0.05) | -$29,000  (0.14) | -$23,000  (0.23) |
| Drug price reduced to 10% of original ($5,600) | Societal | -$19,000  (0.08) | -$12,000  (0.26) | -$6,000  (0.42) | $1,000  (0.53) | $8,000†  (0.58) |
|  | HCS | -$18,000  (0.01) | -$13,000  (0.15) | -$7,000  (0.35) | -$1,000  (0.49) | $5,000†  (0.57) |
| Exclude Caregiver QALYs | Societal | -$94,000  (0) | -$88,000  (0) | -$82,000  (0) | -$76,000  (0.03) | -$69,000  (0.06) |
| Using ICER’s background medical cost multipliers | Societal | -$92,000  (0) | -$84,000  (0) | -$77,000  (0.02) | -$69,000  (0.05) | -$62,000  (0.11) |
|  | HCS | -$91,000  (0) | -$85,000  (0) | -$78,000  (0) | -$72,000  (0.02) | -$66,000  (0.05) |
| Excluding treatment time and transportation costs | Societal | -$86,000  (0) | -$79,000  (0) | -$72,000  (0.02) | -$65,000  (0.06) | -$57,000  (0.13) |
| Caregiver time valued at home health aid wage | Societal | -$94,000  (0) | -$87,000  (0) | -$79,000  (0.01) | -$72,000  (0.05) | -$65,000  (0.1) |
| Using ICER’s productivity approach | Societal | -$96,000  (0) | -$89,000  (0) | -$82,000  (0) | -$74,000  (0.03) | -$67,000  (0.08) |

Abbreviations: r – Discount Rate, INMB – Incremental Net Monetary Benefit, QALY – Quality-Adjusted Life Year, WTP – Willingness to Pay, Societal – Societal Perspective, HCS – Healthcare System, SD – Standard Deviation

\# Values are rounded to the nearest $1,000.

† Positive values indicate treatment is optimal

eFigure 3: Expected Incremental QALYs and Costs vs. MCI-Specific HR

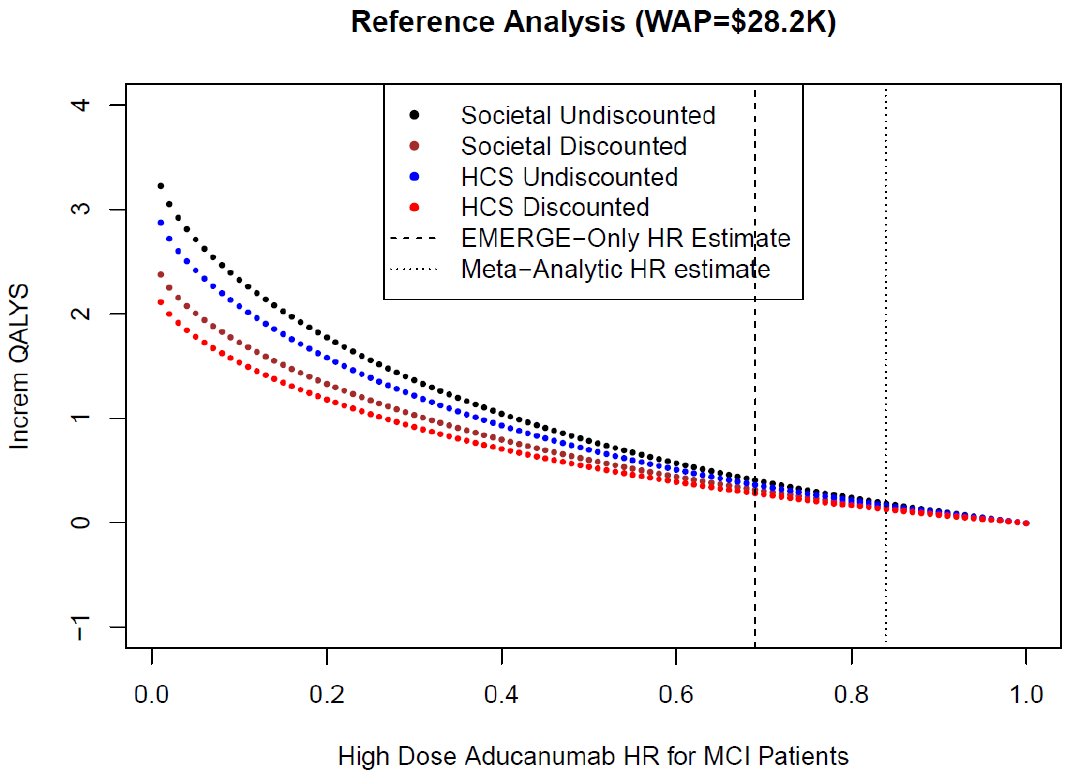

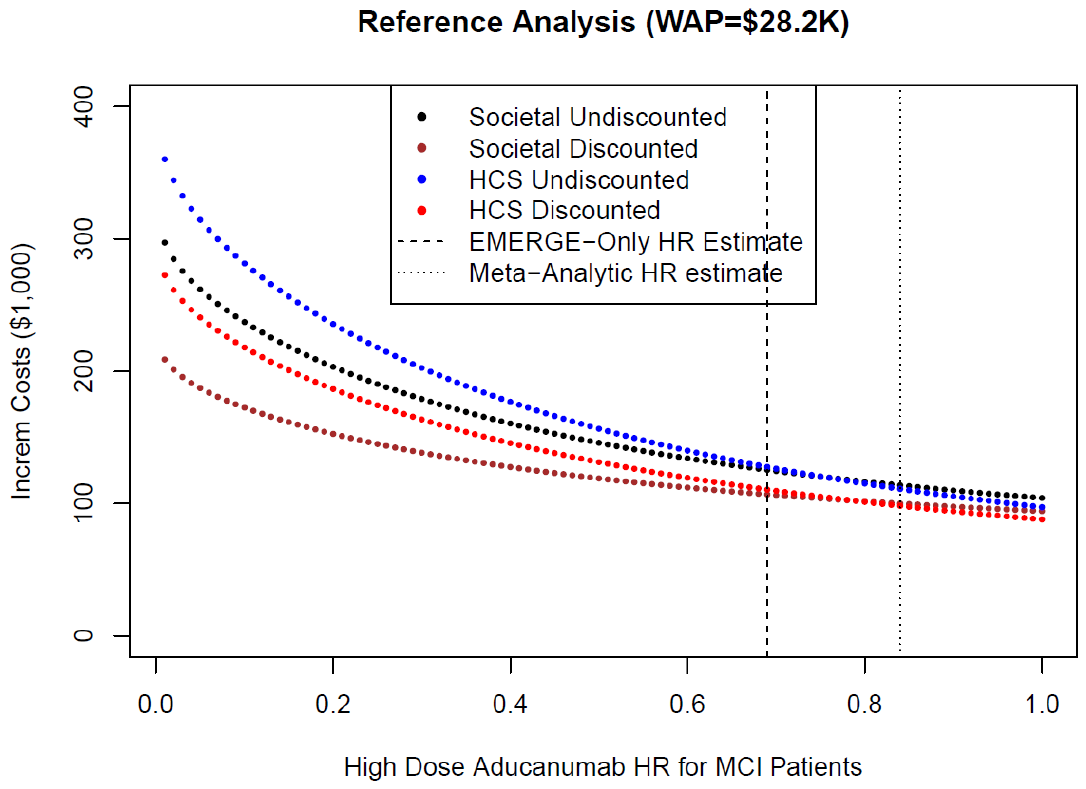

Abbreviations: MCI – Mild Cognitive Impairment, HR – Hazard Ratio, WAP – Wholesale Acquisition Price, K – Thousand, WTP – Willingness to Pay

eFigure 3 depicts how the posterior mean incremental (treatment minus control) QALYs (top) and Costs (bottom), vary with the true MCI-specific HR value for high-dose aducanumab. Results are presented for the societal and healthcare perspectives, discounted (3%) and undiscounted.

eFigure 4: Expected INMB vs. MCI-Specific HR for Reference Analysis (WAP=$28K) and WAP=$5.6K

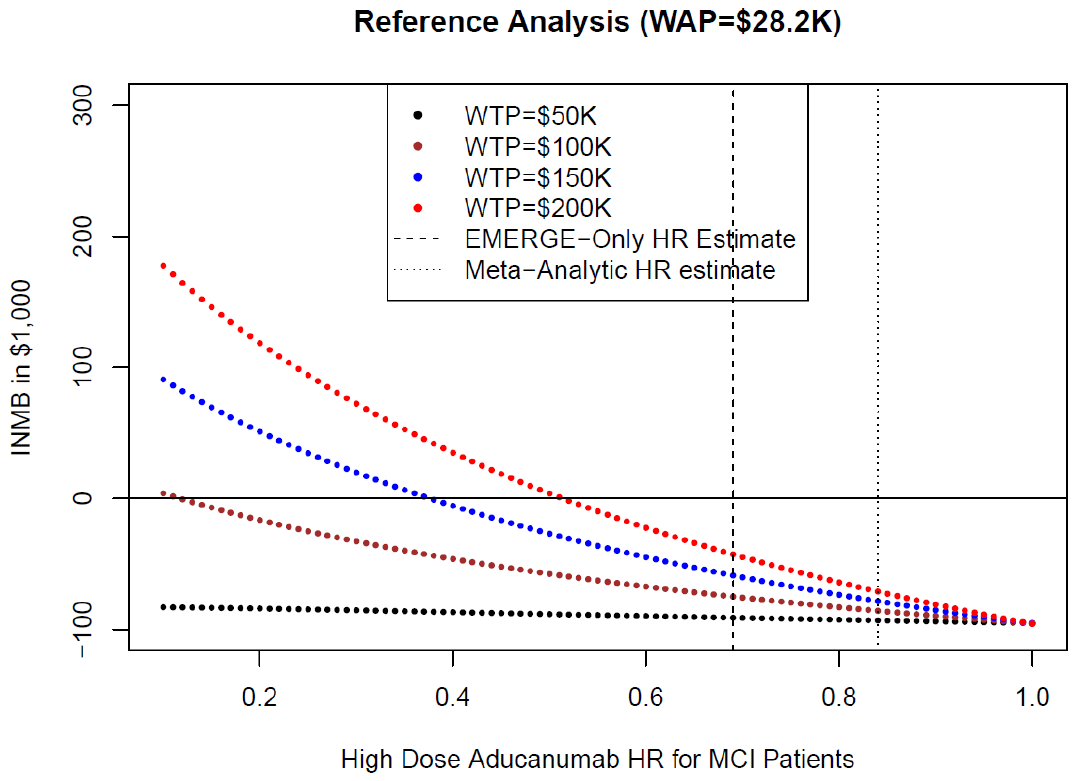

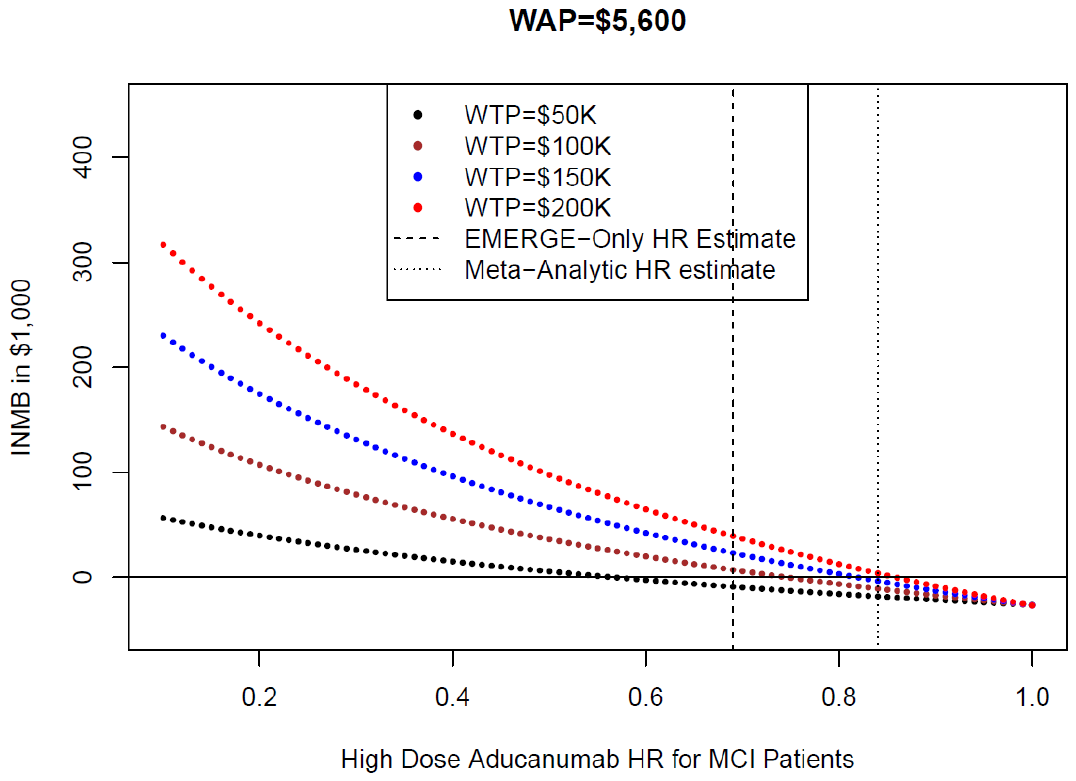

Abbreviations: MCI – Mild Cognitive Impairment, HR – Hazard Ratio, WAP – Wholesale Acquisition Price, K – Thousand, WTP – Willingness to Pay, INMB – Incremental Net Monetary Benefit

eFigure 4 depicts how the posterior mean INMB varies with the true MCI-specific HR value for high-dose aducanumab. Results are presented for the current WAP (top) and WAP=$5.6k (bottom) - 10% of the original manufacturer-proposed price. In both cases, results are given for WTP values between $50K and 200K and were calculated from the societal perspective with 3% discounting. The horizontal line at INMB=0 represents the threshold at which (above) treatment is preferred.

eTable 8: Range of MCI-Specific HR Values at Which High-Dose Aducanumab is Preferred to Usual Care

| WTP | Reference Analysis  (WAP=$28.2K) | WAP=$14.1K | WAP=$5.6K |
| --- | --- | --- | --- |
| $50,000 | -- | ≤0.13 | ≤0.56 |
| $100,000 | ≤0.11 | ≤0.50 | ≤0.74 |
| $150,000 | ≤0.37 | ≤0.64 | ≤0.81 |
| $200,000 | ≤0.51 | ≤0.72 | ≤0.85 |
| $250,000 | ≤0.60 | ≤0.77 | ≤0.88 |
| $300,000 | ≤0.66 | ≤0.80 | ≤0.89 |
| $350,000 | ≤0.70 | ≤0.83 | ≤0.91 |
| $400,000 | ≤0.73 | ≤0.85 | ≤0.92 |
| $450,000 | ≤0.76 | ≤0.86 | ≤0.93 |
| $500,000 | ≤0.78 | ≤0.87 | ≤0.93 |

Abbreviations: MCI – Mild Cognitive Impairment, HR – Hazard Ratio, WTP – Willingness to Pay (for a QALY), WAP – Wholesale Acquisition Price, K – Thousand

eTable 8 gives the threshold MCI-specific HR values (for high-dose aducanumab) at which (and for lower values) treatment would be an efficient use of resources by the criterion of INMB>0. Threshold values are given for a range of WTP values and the 3 WAP scenarios. For example, at a WTP of $150K, if the WAP was $14.1K (after another 50% reduction), the true MCI-specific HR would have to be less than or equal to 0.64 for treatment to be cost-effective. Results were calculated from the societal perspective with 3% discounting.

*Value of a Future Trial*

eTable 9: Estimated Expected Direct Research Costs Associated with an RCT Evaluating Aducanumab

| Total Trial N | Expected Direct Research Costs#  (million dollars) | | | | |
| --- | --- | --- | --- | --- | --- |
|  | Trial Follow Up Length (months) | | | | |
|  | 12 | 18 | 24 | 30 | 36 |
| 1,000 | 19.1  (13.7, 28.5) | 26.5  (19, 39.6) | 33.9  (24.3, 50.6) | 41.1  (29.5, 61.4) | 48.3  (34.6, 72.1) |
| 1,500 | 28.6  (20.5, 42.7) | 39.8  (28.5, 59.3) | 50.9  (36.5, 75.9) | 61.7  (44.2, 92) | 72.5  (51.9, 108.2) |
| 2,000 | 38.2  (27.4, 57) | 53  (38, 79.1) | 67.9  (48.6, 101.2) | 82.3  (58.9, 122.7) | 96.7  (69.2, 144.2) |
| 2,500 | 47.7  (34.2, 71.2) | 66.3  (47.5, 98.9) | 84.8  (60.8, 126.5) | 102.8  (73.7, 153.4) | 120.8  (86.6, 180.3) |
| 3,000 | 57.3  (41, 85.5) | 79.5  (57, 118.7) | 101.8  (72.9, 151.9) | 123.4  (88.4, 184.1) | 145  (103.9, 216.3) |
| 3,500 | 66.8  (47.9, 99.7) | 92.8  (66.5, 138.4) | 118.8  (85.1, 177.2) | 144  (103.1, 214.8) | 169.2  (121.2, 252.4) |
| 4,000 | 76.4  (54.7, 114) | 106.1  (76, 158.2) | 135.7  (97.2, 202.5) | 164.5  (117.8, 245.4) | 193.3  (138.5, 288.4) |
| 4,500 | 85.9  (61.6, 128.2) | 119.3  (85.5, 178) | 152.7  (109.4, 227.8) | 185.1  (132.6, 276.1) | 217.5  (155.8, 324.5) |
| 5,000 | 95.5  (68.4, 142.5) | 132.6  (95, 197.8) | 169.6  (121.5, 253.1) | 205.7  (147.3, 306.8) | 241.7  (173.1, 360.5) |
| 5,500 | 105  (75.2, 156.7) | 145.8  (104.5, 217.5) | 186.6  (133.7, 278.4) | 226.2  (162, 337.5) | 265.8  (190.4, 396.6) |
| 6,000 | 114.6  (82.1, 170.9) | 159.1  (114, 237.3) | 203.6  (145.8, 303.7) | 246.8  (176.8, 368.2) | 290  (207.7, 432.6) |
| 6,500 | 124.1  (88.9, 185.2) | 172.3  (123.4, 257.1) | 220.5  (158, 329) | 267.3  (191.5, 398.8) | 314.1  (225, 468.7) |
| 7,000 | 133.7  (95.8, 199.4) | 185.6  (132.9, 276.9) | 237.5  (170.1, 354.3) | 287.9  (206.2, 429.5) | 338.3  (242.3, 504.7) |
| 7,500 | 143.2  (102.6, 213.7) | 198.9  (142.4, 296.7) | 254.5  (182.3, 379.6) | 308.5  (221, 460.2) | 362.5  (259.7, 540.8) |
| 8,000 | 152.8  (109.4, 227.9) | 212.1  (151.9, 316.4) | 271.4  (194.4, 404.9) | 329  (235.7, 490.9) | 386.6  (277, 576.8) |
| 8,500 | 162.3  (116.3, 242.2) | 225.4  (161.4, 336.2) | 288.4  (206.6, 430.3) | 349.6  (250.4, 521.6) | 410.8  (294.3, 612.9) |
| 9,000 | 171.9  (123.1, 256.4) | 238.6  (170.9, 356) | 305.4  (218.7, 455.6) | 370.2  (265.2, 552.2) | 435  (311.6, 648.9) |
| 9,500 | 181.4  (130, 270.7) | 251.9  (180.4, 375.8) | 322.3  (230.9, 480.9) | 390.7  (279.9, 582.9) | 459.1  (328.9, 685) |
| 10,000 | 191  (136.8, 284.9) | 265.1  (189.9, 395.5) | 339.3  (243, 506.2) | 411.3  (294.6, 613.6) | 483.3  (346.2, 721) |

Abbreviations: RCT – Randomized Control Trial, N – Sample Size

#Primary values rely on an estimate of the median unit cost, i.e., a visit to a research clinic. Values based on the lower and upper quartile unit costs are given in parentheses below. Costs are discounted at 3%, and values are rounded to the nearest $100,000

eTable 10: Estimated Expected Opportunity Costs of Conducting an RCT Evaluating Aducanumab

| Total Trial N | CMS Initial Coverage Decision# | Expected Opportunity Costs†  (million dollars) | | | | | |
| --- | --- | --- | --- | --- | --- | --- | --- |
|  |  | WTP ($1,000) | | | | | |
|  |  | 50 | 100 | 150 | 200 | 250 | 300 |
| 1,000 | Cover Treatment | -45.3 | -42.1 | -38.8 | -35.5 | -32.2 | -28.9 |
|  | Don’t Cover | 45.3 | 42.1 | 38.8 | 35.5 | 32.2 | 28.9 |
| 1,500 | Cover Treatment | -68 | -63.1 | -58.1 | -53.2 | -48.3 | -43.3 |
|  | Don’t Cover | 68 | 63.1 | 58.1 | 53.2 | 48.3 | 43.3 |
| 2,000 | Cover Treatment | -90.7 | -84.1 | -77.5 | -70.9 | -64.4 | -57.8 |
|  | Don’t Cover | 90.7 | 84.1 | 77.5 | 70.9 | 64.4 | 57.8 |
| 2,500 | Cover Treatment | -113.4 | -105.1 | -96.9 | -88.7 | -80.5 | -72.2 |
|  | Don’t Cover | 113.4 | 105.1 | 96.9 | 88.7 | 80.5 | 72.2 |
| 3,000 | Cover Treatment | -136 | -126.2 | -116.3 | -106.4 | -96.5 | -86.7 |
|  | Don’t Cover | 136 | 126.2 | 116.3 | 106.4 | 96.5 | 86.7 |
| 3,500 | Cover Treatment | -158.7 | -147.2 | -135.7 | -124.2 | -112.6 | -101.1 |
|  | Don’t Cover | 158.7 | 147.2 | 135.7 | 124.2 | 112.6 | 101.1 |
| 4,000 | Cover Treatment | -181.4 | -168.2 | -155.1 | -141.9 | -128.7 | -115.6 |
|  | Don’t Cover | 181.4 | 168.2 | 155.1 | 141.9 | 128.7 | 115.6 |
| 4,500 | Cover Treatment | -204.1 | -189.3 | -174.4 | -159.6 | -144.8 | -130 |
|  | Don’t Cover | 204.1 | 189.3 | 174.4 | 159.6 | 144.8 | 130 |
| 5,000 | Cover Treatment | -226.7 | -210.3 | -193.8 | -177.4 | -160.9 | -144.5 |
|  | Don’t Cover | 226.7 | 210.3 | 193.8 | 177.4 | 160.9 | 144.5 |
| 5,500 | Cover Treatment | -249.4 | -231.3 | -213.2 | -195.1 | -177 | -158.9 |
|  | Don’t Cover | 249.4 | 231.3 | 213.2 | 195.1 | 177 | 158.9 |
| 6,000 | Cover Treatment | -272.1 | -252.3 | -232.6 | -212.8 | -193.1 | -173.3 |
|  | Don’t Cover | 272.1 | 252.3 | 232.6 | 212.8 | 193.1 | 173.3 |
| 6,500 | Cover Treatment | -294.8 | -273.4 | -252 | -230.6 | -209.2 | -187.8 |
|  | Don’t Cover | 294.8 | 273.4 | 252 | 230.6 | 209.2 | 187.8 |
| 7,000 | Cover Treatment | -317.4 | -294.4 | -271.4 | -248.3 | -225.3 | -202.2 |
|  | Don’t Cover | 317.4 | 294.4 | 271.4 | 248.3 | 225.3 | 202.2 |
| 7,500 | Cover Treatment | -340.1 | -315.4 | -290.7 | -266.1 | -241.4 | -216.7 |
|  | Don’t Cover | 340.1 | 315.4 | 290.7 | 266.1 | 241.4 | 216.7 |
| 8,000 | Cover Treatment | -362.8 | -336.5 | -310.1 | -283.8 | -257.5 | -231.1 |
|  | Don’t Cover | 362.8 | 336.5 | 310.1 | 283.8 | 257.5 | 231.1 |
| 8,500 | Cover Treatment | -385.5 | -357.5 | -329.5 | -301.5 | -273.6 | -245.6 |
|  | Don’t Cover | 385.5 | 357.5 | 329.5 | 301.5 | 273.6 | 245.6 |
| 9,000 | Cover Treatment | -408.1 | -378.5 | -348.9 | -319.3 | -289.6 | -260 |
|  | Don’t Cover | 408.1 | 378.5 | 348.9 | 319.3 | 289.6 | 260 |
| 9,500 | Cover Treatment | -430.8 | -399.5 | -368.3 | -337 | -305.7 | -274.5 |
|  | Don’t Cover | 430.8 | 399.5 | 368.3 | 337 | 305.7 | 274.5 |
| 10,000 | Cover Treatment | -453.5 | -420.6 | -387.7 | -354.7 | -321.8 | -288.9 |
|  | Don’t Cover | 453.5 | 420.6 | 387.7 | 354.7 | 321.8 | 288.9 |

Abbreviations: RCT – Randomized Control Trial, WTP – Willingness to Pay (for a QALY)

\# Cover Treatment signifies (counterfactually) CMS’s initial decision is to cover aducanumab while Don’t Cover means CMS’s (actual) initial decision is to not cover aducanumab (outside of possibly research studies, e.g. CED).

†The opportunity costs of conducting a trial include treating half the enrolled participants (assuming a balance trial) with the non-standard treatment. For example, if CMS’s had initially decided to cover aducanumab, the opportunity cost of the trial would have been the loss from treating half of the trial participants without aducanumab. Cost savings are possible if the standard treatment is not optimal. However, values are given as costs meaning a positive value represents a real cost while a negative value represents a cost-saving. Opportunity costs are calculated based on the reference case analysis and from the societal perspective, with benefits and costs discounted at 3%. Values are rounded to the nearest $100,000.

eFigure 5: Population EVSI, Trial Costs, and ENGS (WTP=$150K) Given CMS’s Do Not Cover Decision

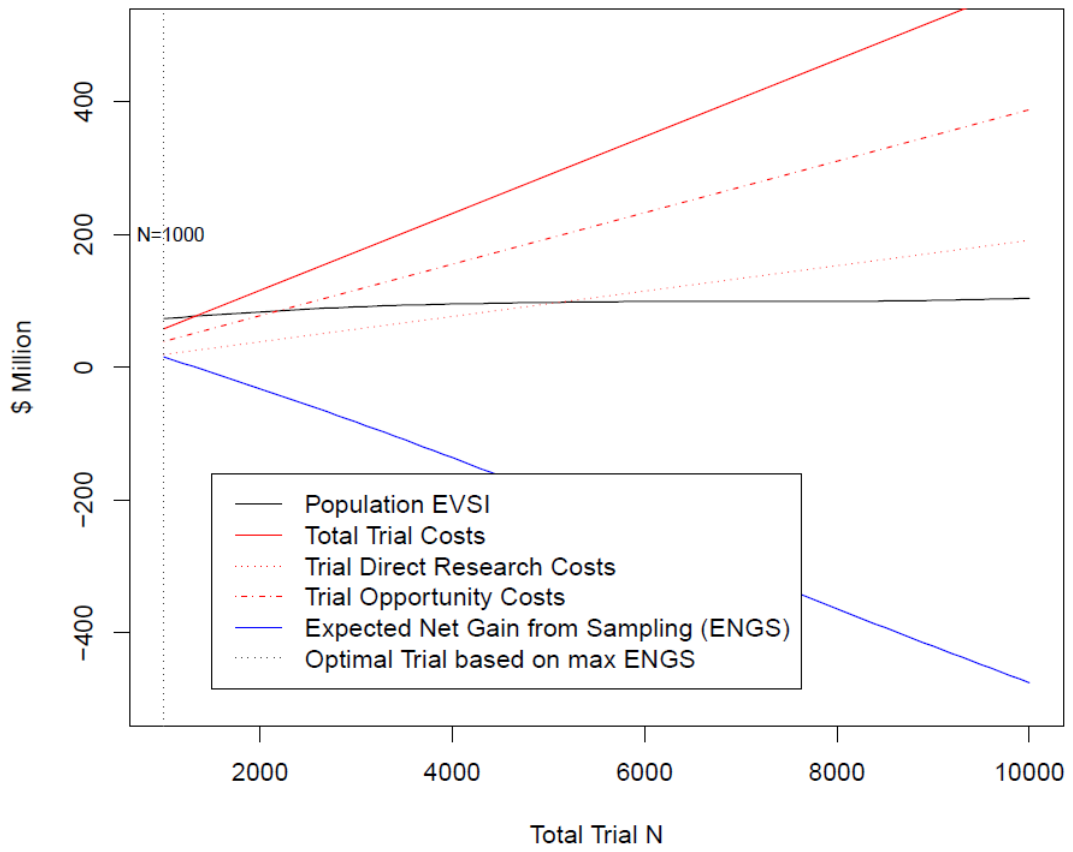

Abbreviations: EVSI – Expected Value of Sample Information, ENGS – Expected Net Gain from Sampling, WTP – Willingness to Pay (for a quality-adjusted life year), CMS – Center for Medicare and Medicaid Services, N – Sample Size

eFigure 5 displays the population EVSI, the expected costs (direct, opportunity, and total), and the ENGS of a future RCT with 12 months follow up as a function of the trial’s sample size. Population EVSI, opportunity costs, and ENGS are calculated given CMS’s actual initial decision not to cover aducanumab for the general population but assuming it will revisit its decision after trial results become available and will make its post-trial decision on efficiency grounds, i.e., based on the sign of INMB. Furthermore, population EVSI, opportunity costs, and ENGS are calculated using the reference-case analysis, taking a societal perspective, assuming a WTP of $150,000 per QALY, and with costs and health effects discounted at 3%. Expected direct research costs are calculated using the median unit cost (our baseline assumption).

The population-EVSI increases slightly with increasing sample size while total costs increase notably. The ENGS thus decreases monotonically across considered sample sizes and is negative at sample sizes >1,500. Thus the smallest considered trial (N=1,000) represents the optimal trial. The choice of unit cost (low, median, or high) had no effect on this result. Notably, if CMS were to make its post-trial decision based on the criterion of statistical significance, the population EVSI would become large negative but large in absolute magnitude, decreasing monotonically from -$25B to -$50B (billion) as sample size increases. This reflects the increasing chance (due to increasing statistical power) that a trial would lead CMS to change its decision and cover aducanumab. Thus no trial would be optimal in this context.

eFigure 6: Population EVSI, Trial Costs, and ENGS (WTP=$200K) Given CMS Did Not Cover

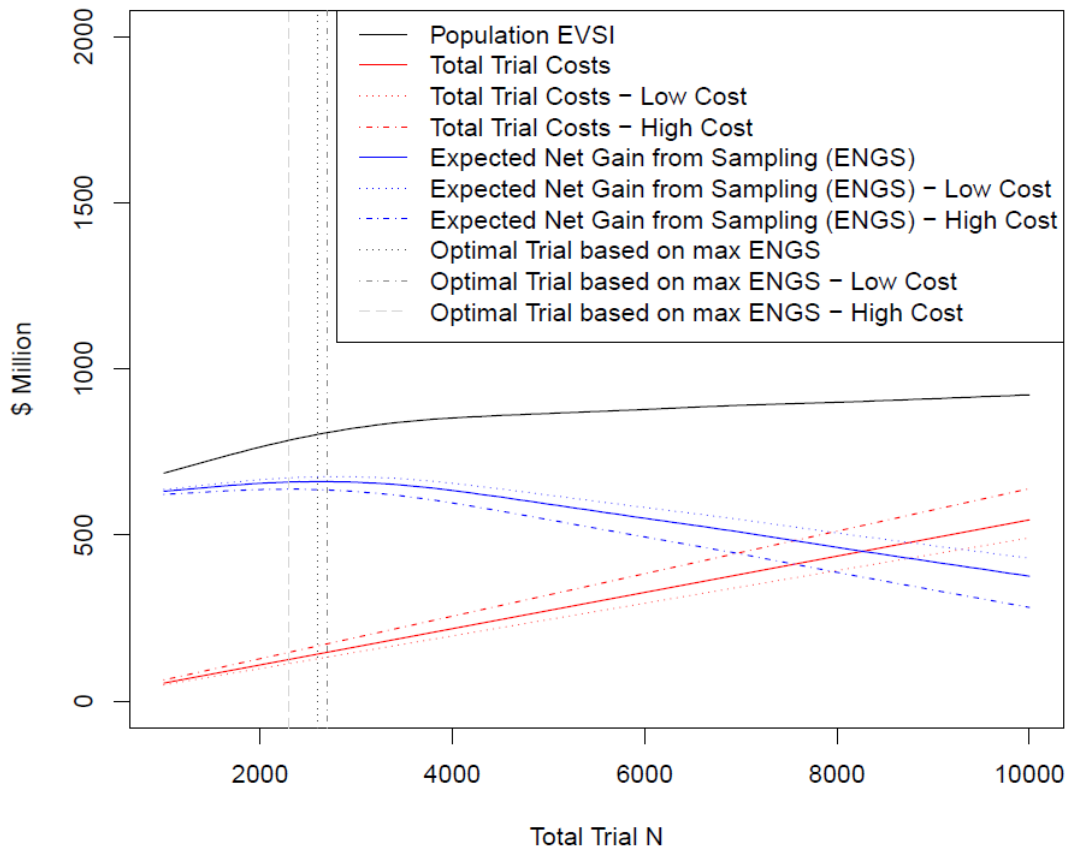

Abbreviations: EVSI – Expected Value of Sample Information, ENGS – Expected Net Gain from Sampling, WTP – Willingness to Pay (for a quality-adjusted life year), CMS – Center for Medicare and Medicaid Services, N – Sample Size

eFigure 6 displays the population EVSI, the total expected costs, and the ENGS of a future RCT with 12 months follow up as a function of the trial’s sample size. Population EVSI, opportunity costs, and ENGS are calculated given CMS’s initial decision not to cover aducanumab (other than possibly in a research setting) but assuming it will revisit its decision after trial results become available and will make its post-trial decision on efficiency grounds, i.e., based on the sign of INMB. Furthermore, population EVSI, opportunity costs, and ENGS are calculated using the reference-case analysis, taking a societal perspective, assuming a WTP of $200,000 per QALY, and with costs and health effects discounted at 3%. In eFigure 6, we demonstrate the effect of different direct research unit cost assumptions on total trial costs and ENGS. Both total trial costs and ENGS are given for the three unit cost assumptions – median (unnamed), low cost, and high cost, and the optimal trial sample size is identified in each case. The population-EVSI increases modestly (with a decreasing rate) with increasing sample size while total costs increase notably. The ENGS increase modestly up to a sample size around 2,000 and then decreases monotonically and more significantly with increasing sample sizes. The optimal trial sample size varies somewhat with the unit cost assumption ranging from 1,700 to 2,100 with the high and low unit cost assumptions, respectively. Again, if CMS were to make its post-trial decision based on the criterion of statistical significance, the population EVSI would be negative and increases dramatically in absolute magnitude, decreasing monotonically from -$25B to -$50B as sample size increases. This reflects the increasing chance (due to increasing statistical power) that a trial would lead CMS to change its decision and cover aducanumab.

eFigure 7: Population EVSI, Trial Costs, and ENGS (WTP=$100K) Had CMS Covered Aducanumab

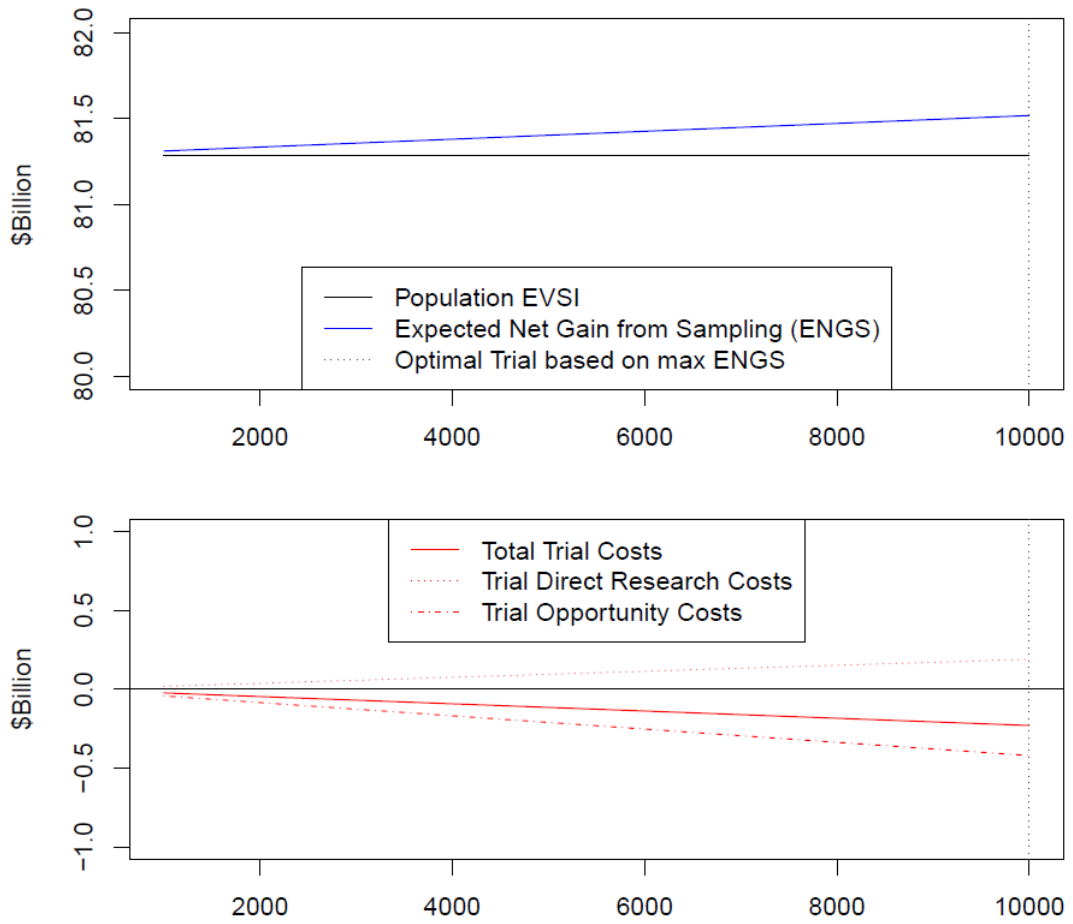

Abbreviations: EVSI – Expected Value of Sample Information, ENGS – Expected Net Gain from Sampling, WTP – Willingness to Pay (for a quality-adjusted life year), CMS – Center for Medicare and Medicaid Services, N – Sample Size

eFigure 7 displays the population EVSI, the expected costs (direct research, opportunity, and total), and the ENGS of a future RCT with 12 months follow up as a function of the trial’s sample size. Population EVSI, opportunity costs, and ENGS are calculated under the counterfactual scenario where CMS makes an initial decision to cover aducanumab but then will revisit its decision after trial results become available and will make its post-trial decision on efficiency grounds, i.e., based on the sign of INMB. Furthermore, population EVSI, opportunity costs, and ENGS are calculated using the reference-case analysis, taking a societal perspective, assuming a WTP of $100,000 per QALY, and with costs and health effects discounted at 3%. Expected direct research costs are calculated using the median unit cost (our baseline assumption). The population-EVSI increases slightly with increasing sample size. While direct research costs increase with sample size, total trial costs decrease monotonically and are negative (implying cost-savings) over all considered sample sizes. This is due to negative opportunity costs (cost-savings) associated with not giving half of trial participants the standard treatment (aducanumab) that increase in absolute magnitude as N increases. Thus, somewhat counterintuitively, ENGS increases monotonically over the considered sample sizes and the largest possible trial (10,000) is the optimal trial. The choice of unit cost (low, median, or high) had no effect on this result. If CMS were to make its post-trial decision based on the criterion of statistical significance, the population EVSI would remain large and positive but would decrease in magnitude. This is because, although the trial would still offer CMS the opportunity to change its costly initial decision and decide not to cover aducanumab, with larger sample sizes, the power of such a trial would increase making it less likely this reversal would happen.

eFigure 8: Population EVSI, Trial Costs, and ENGS (WTP=$150K) Had CMS Covered Aducanumab

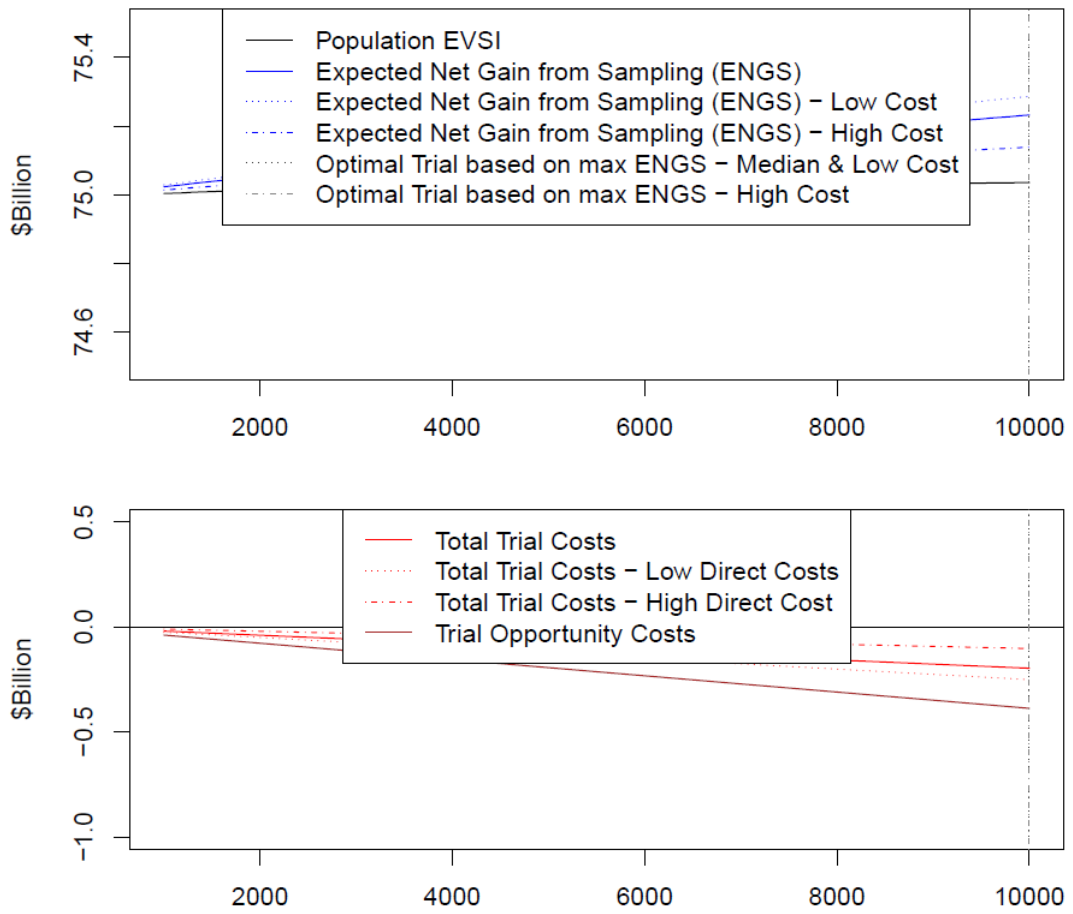

Abbreviations: EVSI – Expected Value of Sample Information, ENGS – Expected Net Gain from Sampling, WTP – Willingness to Pay (for a quality-adjusted life year), CMS – Center for Medicare and Medicaid Services, N – Sample Size

eFigure 8 displays the population EVSI, the total expected costs, and the ENGS of a future RCT with 12 months follow up as a function of the trial’s sample size. Population EVSI, opportunity costs, and ENGS are calculated assuming (counterfactually) CMS made an initial decision to cover aducanumab but then will revisit its decision after trial results become available and will make its post-trial decision on efficiency grounds, i.e., based on the sign of INMB. Furthermore, population EVSI, opportunity costs, and ENGS are calculated using the reference-case analysis, taking a societal perspective, assuming a WTP of $150,000 per QALY, and with costs and health effects discounted at 3%. In eFigure 8, we demonstrate the effect of different unit cost assumptions (for direct research) on total trial costs and ENGS. Both total trial costs and ENGS are given for the 3 unit cost assumptions – median (unnamed), low cost, and high cost - and the optimal trial sample size is identified in each case. The population-EVSI increases slightly with increasing N. Using the low and median unit cost assumptions, total costs are negative and decrease montonically with larger N due to increasing cost-savings from not treating half of trial participants with aducanumab. Thus under these cost assumptions, the optimal trial is the largest – 10,000. However, under the high unit cost assumption, the direct costs of research slightly outweigh the cost-savings and do so increasingly as N increases, and thus total costs increases monotonically. The composition of these different effects leads ENGS (under the high cost assumption) to increase at first and peak at N=6,200 and then decrease. If CMS were to make its post-trial decision based on the criterion of statistical significance, the population EVSI would remain large and positive but would be smaller than if CMS made its post-trial decision based on efficiency. This is because the trial would still offer CMS the opportunity to change its costly initial decision and decide not to cover aducanumab. However, with larger sample sizes, the power would increase making it less likely this reversal would happen.

eTable 11: Ranking of Decision Alternatives and Expected Opportunity Loss (WTP>300,000)

| Post-Trial Decision Criterion | Efficiency  (INMB>0) | | Statistical Significance  (P<0.05) | |
| --- | --- | --- | --- | --- |
| Trial Design | Max. ENGS  (2.a.i/4.a.i)# | ENVISION-Type  Trial (2.a.ii/4.a.ii)† | Max. ENGS  (2.b.i/4.b.i)# | ENVISION-Type Trial (2.b.ii/4.b.ii)† |
| WTP††  ($1,000) | Ranking (Expected Opportunity Loss of selecting a suboptimal decision given as forgone $Billions of NMB*) of Each Decision Alternative.¶ | | | |
| 350 | 1.NoTrt/Trial  2.NoTrt/NoTrial (8.76)  3.Trt/Trial (11.49)  4.Trt/NoTrial (70.97) | 1.NoTrt/Trial  2.NoTrt/NoTrial (7.73)  3.Trt/Trial (14.62)  4.Trt/NoTrial (69.94) | 1.NoTrt/NoTrial  2.NoTrt/Trial (9.25)  3.Trt/Trial (22.38)  4.Trt/NoTrial (62.21) | 1.NoTrt/NoTrial  2.NoTrt/Trial (13.39)  3.Trt/Trial (29.14)  4.Trt/NoTrial (62.21) |
| 400 | 1.NoTrt/Trial  2.Trt/Trial (9.72)  3.NoTrt/NoTrial(13.19)  4.Trt/NoTrial (67.40) | 1.NoTrt/Trial  2.NoTrt/NoTrial(11.62)  3.Trt/Trial (12.48)  4.Trt/NoTrial (65.83) | 1.NoTrt/NoTrial  2.NoTrt/Trial (5.32)  3.Trt/Trial (16.61)  4.Trt/NoTrial (54.21) | 1.NoTrt/NoTrial  2.NoTrt/Trial (8.88)  3.Trt/Trial (22.45)  4.Trt/NoTrial (54.21) |
| 450 | 1.NoTrt/Trial  2.Trt/Trial (7.94)  3.NoTrt/NoTrial(18.23)  4.Trt/NoTrial (64.44) | 1.NoTrt/Trial  2.Trt/Trial (10.31)  3.NoTrt/NoTrial(16.11)  4.Trt/NoTrial (62.32) | 1.NoTrt/NoTrial  2.NoTrt/Trial (1.33)  2.Trt/Trial (10.79)  3.Trt/NoTrial (46.21) | 1.NoTrt/NoTrial  2.NoTrt/Trial (4.36)  3.Trt/Trial (15.76)  4.Trt/NoTrial (46.21) |
| 500 | 1.NoTrt/Trial  2.Trt/Trial (6.11)  3.NoTrt/NoTrial(23.77)  4.Trt/NoTrial (61.98) | 1.NoTrt/Trial  2.Trt/Trial (8.11)  3.NoTrt/NoTrial(21.13)  4.Trt/NoTrial (59.35) | 1.NoTrt/Trial  2.NoTrt/NoTrial(2.73)  3.Trt/Trial (7.63)  4.Trt/NoTrial (40.94) | 1.NoTrt/Trial  2.NoTrt/NoTrial(0.16)  3.Trt/Trial (9.22)  4.Trt/NoTrial (38.37) |

Abbreviations: NMB – Net Monetary Benefit, INMB – Incremental NMB, ENGS – Expected Net Gain from Sampling, WTP – Willingness to Pay, NoTrt – Do Not Cover Aducanumab, Trt – Cover Aducanumab, NoTrial – Do Not Perform Trial, Trial – Perform Trial.

#In these analyses (columns 2 and 4), in each cell, decision alternatives including a trial are assumed to include the optimally designed trial, where optimality is defined in terms of ENGS and dependent on the WTP for a QALY and CMS’s initial coverage decision and assumed post-trial decision criterion. In cases where the optimal trial is actually no trial, i.e., no design has positive ENGS, the least bad trial is used (smallest loss).

†In these analysis, any decision alternative that includes a trial, e.g., Trt/Trial, is assumed to include a trial designed to have at least 90% power to detect the effect size reported in the EMERGE trial given 18 months of follow-up (N≈1,400). Such a trial is comparable to the upcoming phase-4 trial (‘ENVISION’) planned by the manufacturer.

*Expected opportunity losses represent forgone NMB (rounded to the nearest $10,000,000) associated with selecting each option instead of the best.

¶ Analyses are done from the societal perspective, using the reference-case analysis, with costs and health effects discounted at 3%. Expected direct research costs were calculated using the median unit cost (our baseline assumption).

††Results for WTP≤$300K are given in Table 3.

eTable 12: Impact of Modeling Scenarios on Decision Alternative Rankings

| Modeling Scenario or Assumption | Ranking (Expected Opportunity Loss of selecting a suboptimal decision given as forgone $Billions of NMB*) of Each Decision Alternative Assuming an ENVISION-like Trial† and a Post-Trial Decision Criterion of Efficiency (INMB>0)¶ | | | |
| --- | --- | --- | --- | --- |
|  | WTP ($1,000) | | | |
|  | 50 | 100 | 150 | 200 |
| Reference Analysis | 1.NoTrt/NoTrial  2.NoTrt/Trial (0.10)  3.Trt/Trial (26.73)  4.Trt/NoTrial (110.20) | 1.NoTrt/NoTrial  2.NoTrt/Trial (0.10)  3.Trt/Trial (24.79)  4.Trt/NoTrial (102.20*)* | 1.NoTrt/NoTrial  2.NoTrt/Trial (0.01)  3.Trt/Trial (22.77)  4.Trt/NoTrial (94.40) | 1.NoTrt/Trial  2.NoTrt/NoTrial (0.65)  3.Trt/Trial (20.79)  4.Trt/NoTrial (86.86) |
| Optimistic Analysis | 1.NoTrt/NoTrial  2.NoTrt/Trial (0.12)  3.Trt/Trial (33.22)  4.Trt/NoTrial (136.97) | 1.NoTrt/NoTrial  2.NoTrt/Trial (0.10)  3.Trt/Trial (26.82)  4.Trt/NoTrial (110.57) | 1.NoTrt/NoTrial  2.NoTrt/Trial (0.09)  3.Trt/Trial (20.43)  4.Trt/NoTrial (84.18) | *1.NoTrt/NoTrial* §  *2.NoTrt/Trial (0.05)*  2.Trt/Trial (14.01)  3.Trt/NoTrial (57.78) |
| WAP of  $14,100 | 1.NoTrt/NoTrial  2.NoTrt/Trial (0.07)  3.Trt/Trial (13.48)  4.Trt/NoTrial (55.48) | *1.NoTrt/Trial* §  *2.NoTrt/NoTrial (0.33)*  3. Trt/Trial (11.46)  4. Trt/NoTrial (47.81) | *1.NoTrt/Trial* §  *2.NoTrt/NoTrial (2.16)*  3.Trt/Trial (9.43)  4.Trt/NoTrial (41.64) | 1.NoTrt/Trial §  *2.Trt/Trial (5.49)*  *3.NoTrt/NoTrial (7.31)*  4. Trt/NoTrial (36.98) |
| WAP of  $5,600 | *1.NoTrt/Trial* §  *2.NoTrt/NoTrial (0.39)*  3.Trt/Trial (5.41)  4.Trt/NoTrial (22.90) | *1.NoTrt/Trial* §  *2.Trt/Trial (3.32)*  *3.NoTrt/NoTrial (3.49)*  4.Trt/NoTrial (17.99) | *1.NoTrt/Trial* §  *2.Trt/Trial (1.11)*  *3.NoTrt/NoTrial (8.71)*  4.Trt/NoTrial (15.22) | *1.Trt/Trial* §  *2.NoTrt/Trial (1.16)*  *3.Trt/NoTrial (14.66)*  *4.NoTrt/NoTrial(16.15)* |

Abbreviations: INMB – Incremental Net Monetary Benefit, WTP – Willingness to Pay, NoTrt – Do Not Cover Aducanumab, Trt – Cover Aducanumab, NoTrial – Do Not Perform Trial, Trial – Perform Trial, WAP – Wholesale Acquisition Price

*Values are rounded to the nearest billion dollars.

†An 18-month follow up with a total sample size of roughly 1,400.

¶ Analyses are done from the societal perspective, using the reference-case analysis, with costs and health effects discounted at 3%. Expected direct research costs were calculated using the median unit cost (our baseline assumption).

§ Indicates the ranking differs from the reference case; relevant alternatives are italicized.

eFigure 9: Power Analysis for a RCT Evaluating the Efficacy of Aducanumab

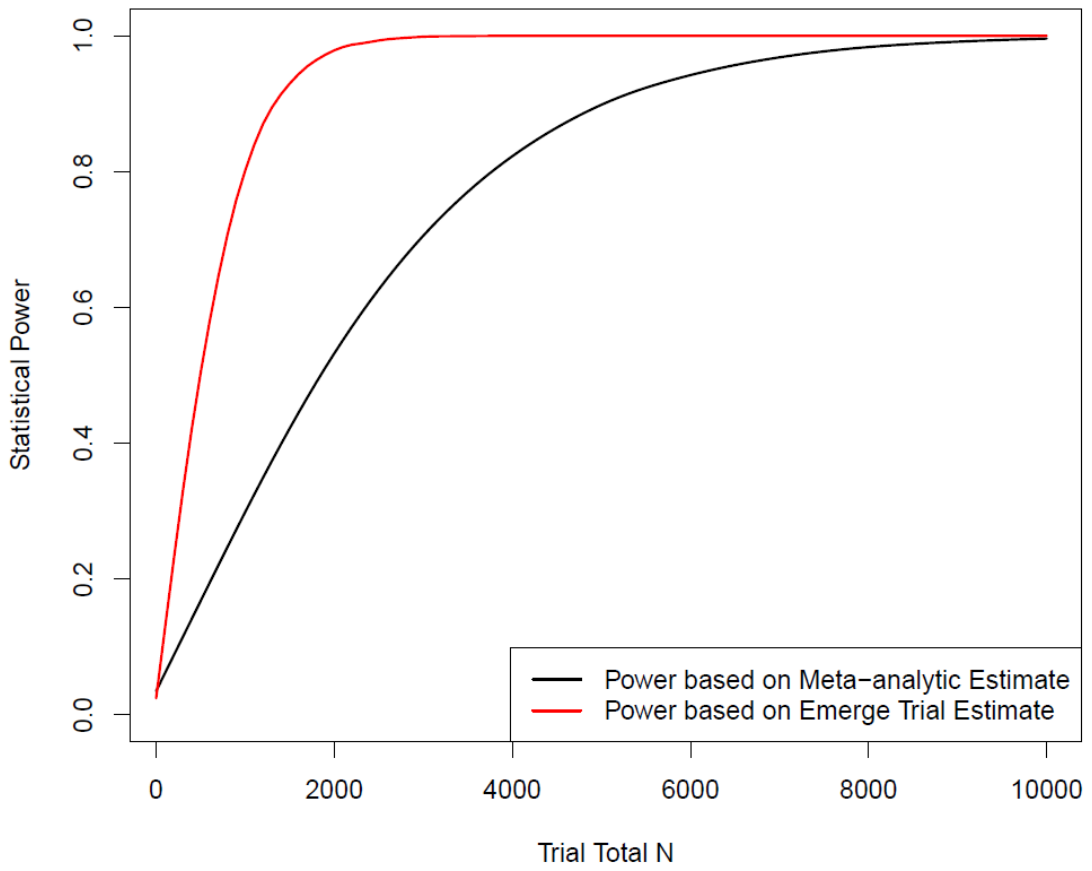

Abbreviations: RCT – Randomized Control Trial, N – Sample Size

eFigure 9 displays the statistical power of a future RCT designed to evaluate the efficacy of aducanumab as a function of the total sample size. We assume the trial would involve 18 months of follow up and the primary endpoint would be time (from MCI) to mild AD with the effect of treatment estimated as a hazard ratio. Two analyses were undertaken, one assuming the meta-analytic HR estimate (0.84) was the true effect and another assuming the EMERGE trial estimate was the true effect (0.69). We assume the latter assumption would be used to design a real trial analogous to the manufacturer’s planned ENVISION trial, but, given current knowledge, we expect the former may give a more realistic power curve. For both analyses, the power curve gives the probability of rejecting the null hypothesis of no effect as a function of sample size given the true effect size and using a two-sided statistical test with a type I error of 0.05. The power analysis was done by simulation study with baseline rates taken from the decision model and allowing for background mortality and attrition. Details are given above in Supplementary methods. To address Monte Carlo error, values are smoothed using penalized cubic splines. If the EMERGE trial estimate were accurate, to achieve 80% or 90% power, a trial would need to randomize 1,000 or 1,325 participants, respectively. For reference, we note that, using only analytic methods, we estimate 229 and 306 transitions to mild AD would need to be observed, respectively. However, if the meta-analytic estimate represented the true effect, 4,271 or 5,743 patients would need to be randomized respectively.

1.	Chhatwal J, Jayasuriya S, Elbasha EH. Changing cycle lengths in state-transition models: challenges and solutions. *Medical Decision Making*. 2016;36(8):952-964.

2.	Jahn B, Kurzthaler C, Chhatwal J, et al. Alternative conversion methods for transition probabilities in state-transition models: validity and impact on comparative effectiveness and cost-effectiveness. *Medical Decision Making*. 2019;39(5):509-522.

3.	Lin L. *Roots of stochastic matrices and fractional matrix powers*. The University of Manchester (United Kingdom); 2011.

4.	Sutton AJ, Abrams KR. Bayesian methods in meta-analysis and evidence synthesis. *Statistical methods in medical research*. 2001;10(4):277-303.

5.	Herring WL, Gould IG, Fillit H, et al. Predicted Lifetime Health Outcomes for Aducanumab in Patients with Early Alzheimer’s Disease. *Neurology and Therapy*. 2021:1-22.

6.	Lin G, Whittington M, Synnott P, et al. *Aducanumab for Alzheimer’s Disease: Effectiveness and Value; Final Evidence Report and Meeting Summary*. 2021. *Institute for Clinical and Economic Review*. August 5, 2021. <https://icer.org/assessment/alzheimers-disease-2021/>

7.	Anderson TS, Ayanian JZ, Souza J, Landon BE. Representativeness of Participants Eligible to Be Enrolled in Clinical Trials of Aducanumab for Alzheimer Disease Compared With Medicare Beneficiaries With Alzheimer Disease and Mild Cognitive Impairment. *JAMA*. 2021;326(16):1627-1629.

8.	Neumann PJ, Sanders GD, Russell LB, Ganiats TG, Siegel JE. *Cost-effectiveness in Health and Medicine*. Oxford University Press; 2017.

9.	Turner RM, Davey J, Clarke MJ, Thompson SG, Higgins JP. Predicting the extent of heterogeneity in meta-analysis, using empirical data from the Cochrane Database of Systematic Reviews. *International journal of epidemiology*. 2012;41(3):818-827.

10.	The Human Mortality Database. Accessed 06/16/2021.

11.	Neumann PJ, Kuntz KM, Leon J, et al. Health utilities in Alzheimer's disease: a cross-sectional study of patients and caregivers. *Medical care*. 1999:27-32.

12.	*Combined FDA and Applicant PCNS Drug Advisory Committee Briefing Document*. 2020.

21.	National Coverage Analysis (NCA) Proposed Decision Memo: Monoclonal Antibodies Directed Against Amyloid for the Treatment of Alzheimer’s Disease (CMS Newsroom)

31.	Physician Fee Scheulde.

32.	Jiao B, Basu A. Catalog of Age- and Medical Condition—Specific Healthcare Costs in the United States to Inform Future Costs Calculations in Cost-Effectiveness Analysis. *Value in Health*. 2021/07/01/ 2021;24(7):957-965. doi:<https://doi.org/10.1016/j.jval.2021.03.006>

33.	Leibson CL, Long KH, Ransom JE, et al. Direct medical costs and source of cost differences across the spectrum of cognitive decline: a population-based study. *Alzheimer's & Dementia*. 2015;11(8):917-932.

34.	Barthold D, Joyce G, Ferido P, et al. Pharmaceutical Treatment for Alzheimer's Disease and Related Dementias: Utilization and Disparities. *J Alzheimers Dis*. 2020;76(2):579-589. doi:10.3233/jad-200133

35.	Administration on Aging. Costs of Care. <https://acl.gov/ltc/costs-and-who-pays/costs-of-care>

36.	Average hourly and weekly earnings of all employees on private nonfarm payrolls by industry sector, seasonally adjusted (Bureau of Labor Statistics, US Department of Labor) (2020).

37.	Occupational Employment and Wages, May 2020 (2020).

38.	Haro J, Kahle-Wrobleski K, Bruno G, et al. Analysis of burden in caregivers of people with Alzheimer’s disease using self-report and supervision hours. *The journal of nutrition, health & aging*. 2014;18(7):677-684.
